# Integrating perturbational screens, eQTL, and GWAS data identifies mediating genes for complex traits

**DOI:** 10.64898/2026.01.05.26343421

**Authors:** Zeyun Lu, Yi Ding, Nathan LaPierre, Lili Wang, Douglas Yao, Nicholas Mancuso, Alexander Gusev

## Abstract

Most current GWAS-eQTL approaches prioritize genes whose mediating effects on complex traits act through *cis*-regulation, while *trans*-acting genes remain largely underexplored. Recent perturbational screening technology provides a novel approach to quantifying *trans*-effects between gene pairs, but its integration with GWAS data remains largely unexamined. We introduce Mr. PEG, a novel framework that integrates perturbational screens, eQTL, and GWAS summary data to identify mediating genes of complex traits. Integrating gene-to-gene effects estimated from perturbational screens and GWAS data across 40 complex traits, Mr. PEG identifies a total of 546 significant mediating genes. These genes are more constrained than background genes and enriched for Gene Ontology terms related to immune response and cellular signaling. Compared to genes identified by GWAS and Mendelian randomization-based approaches, Mr. PEG genes exhibit longer average lengths across enhancers and stronger coexpression. Mr. PEG effects learned from common non-coding variants are associated with rare coding burden effects, highlighting its ability to capture disease-relevant mechanisms missed by approaches focused only on *cis*-eQTLs. We also highlight a case in which Mr. PEG uniquely identifies *PTGS2* as a mediating gene for gout, suggesting potential opportunities for drug repurposing. Our findings demonstrate the value of integrating *trans*-effects informed by experimental perturbation screens and population-scale GWAS and eQTL data to identify disease-relevant mediating genes beyond individual GWAS loci.

## Introduction

A key question in Genome-Wide Association Studies (GWASs) is how genetic variants influence complex traits through intermediate genes and molecular pathways^1,2^. Most significant GWAS associations are in non-coding regions, and are hypothesized to regulate molecular traits such as gene expression, which in turn affect downstream phenotypes^3,4^. Integrative approaches combining GWAS findings with *cis*-expression Quantitative Trait Loci (*cis*-eQTLs), including Mendelian Randomization (MR)^5,6^, Transcriptome-Wide Association Studies (TWASs)^7–11^, and colocalization tests^12,13^, have identified thousands of genes whose predicted mRNA expression is associated with complex traits.

Despite the wide use of GWAS-eQTL integration approaches, recent studies have highlighted differences between these signals^14,15^. Individual *cis*-eQTL associations exhibit systematically different functional properties from GWAS associations, and all predicted *cis*-acting gene expression is estimated to account for only ∼11% of the heritability of complex traits^16^. These findings support conceptual models^17^ where upstream genes regulate the expression of downstream genes (i.e., mediating genes) and, eventually, disease through complex gene regulatory networks^17–20^. Under certain complex trait models, mediating genes are expected to be both more disease-relevant and more difficult to identify through common variant GWAS due to lower statistical power. Ranking genes by their relevance to a trait is essential for understanding the genetic architecture of the trait and for identifying the most trait-relevant mechanisms. However, how to optimally prioritize trait-relevant genes remains a major open question in the field^21^.

Perturbational screens, including Perturb-seq^22^ and CROP-seq^23^, which combines CRISPR screens with single-cell RNA-seq (scRNA-seq), has emerged as a powerful tool for probing gene regulation^22,24^. By measuring transcriptome-wide expression changes following targeted knockouts of specific genes (i.e., perturbed genes), this high-throughput technology enables the characterization of gene-to-gene regulatory effects^25^. Recent approaches have leveraged latent gene modules to enhance statistical power in estimating the effects of genetic perturbations on individual genes^25,26^. Compared with *trans*-eQTL signals, which are often limited by statistical power and can often capture correlational relationships between genes^27^, gene-to-gene effects inferred from CRISPR-based perturbation screens provide more direct evidence of causal *trans*-regulatory relationships rather than association alone^28^. However, few statistical methods currently integrate perturbation-estimated *trans*-effects with GWAS signals to identify trait-specific mediating genes.

In this study, we present a novel statistical framework that unifies *MR*-like tests with *p*erturbational screens, *e*QTL, and *G*WAS summary statistics (Mr. PEG) to identify mediating genes of complex traits. Mr. PEG integrates experimental and population-scale data by modeling SNP-to-trait GWAS signals as the product of three components: (1) SNP-to-gene effect estimated from eQTL studies, (2) gene-to-gene effects estimated from perturbational screens, and (3) gene-to-trait mediating effect, which is the primary parameter of interest (**Fig. 1**). Through extensive simulations, we demonstrate that Mr. PEG is well-calibrated, exhibits high power across realistic parameter spaces, and is robust to model misspecifications. We apply Mr. PEG to 43 GWAS datasets (average effective sample size: 228,500; **Supplementary Table 1**), integrating *cis*-eQTL data from eQTLGen^27^ (n = 31,684) and gene-to-gene effects estimated from perturbation screens (459 perturbed genes on 13,152 downstream genes) measured in macrophage myeloid cell lines^25^. Across 40 traits, Mr. PEG identifies 546 mediating genes, which are more constrained than background genes. We find that these genes exhibit significantly different epigenetic features compared to those identified by GWAS, MR-based methods, and burden tests. Importantly, Mr. PEG effects learned from common non-coding variant GWAS are associated with rare coding burden effects, highlighting its ability to capture disease-relevant genes. We also present examples in which Mr. PEG uniquely identifies mediating genes with potential relevance for drug repurposing in complex diseases. Overall, Mr. PEG provides a powerful framework that integrates experimental perturbation screens and population-scale eQTL and GWAS data to offer novel insights into the genetic architecture of complex traits.

**Fig. 1:**
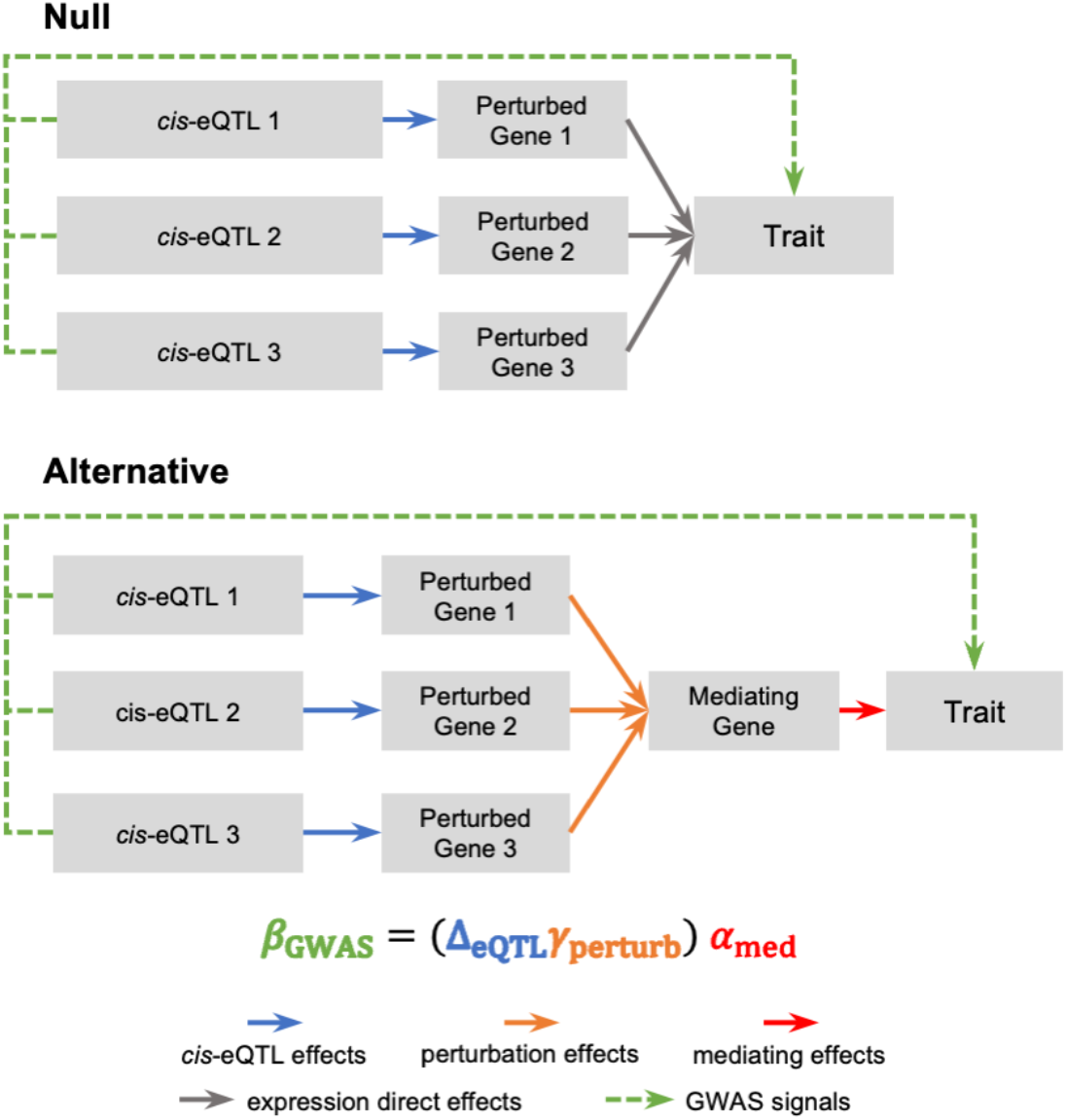
Overview of the Mr. PEG framework. The null assumption of Mr. PEG is that *cis*-eQTLs influence nearby gene expression (Perturbed gene), which in turn affects complex traits, without accounting for gene-to-gene perturbation effects. In contrast, Mr. PEG models complex traits as functions of the expression levels of the mediating genes. The expression level of an mediating gene is further modeled as a function of several perturbed gene expressions, where each perturbed gene is a linear combination of its respective *cis*-region genotypes.

## Results

### Mr. PEG Overview

We provide an overview of the Mr. PEG generative model and its inference framework. In this study, a perturbation refers to a CRISPR-based experimental intervention that knocks out one or more genes (referred to as perturbed genes or upstream genes). Following a perturbation, the expression levels of other genes (referred to as mediating genes or downstream genes) are measured using scRNA-seq or similar techniques. These data can then be analyzed with computational algorithms^25,26^ to infer gene-to-gene effects, that is, how perturbing one gene influences the expression of others.

Mr. PEG estimates the effect size *α* ∈ ℝ of a focal gene’s expression on a complex trait. The expression level of the mediating gene is modeled as a linear combination of the expression levels of perturbed genes, which in turn are modeled as a linear combination of genotyped variants (**Figure 1**). This hierarchical framework captures the relationships between eQTLs, perturbed genes, and mediating genes, ultimately linking genetic variants to complex traits. The mathematical representation is given by

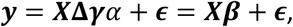

where ***y*** ∈ ℝ^*n*×1^ is the normalized complex trait level measured across *n* individuals with mean of 0 and a standard deviation of 1, ***X*** ∈ ℝ^*n*×*k*^ is the normalized genotype matrix, *k* is the number of eQTLs across *t* perturbed genes, **Δ** ∈ ℝ^*k*×*t*^ represents the eQTL effect sizes on *t* perturbed genes, ***γ*** ∈ ℝ^*t*×1^ denotes the gene-to-gene effect sizes on the mediating gene, *α* ∈ ℝ is the mediating effect size, ***β*** = **Δ*γ****α* ∈ ℝ^*k*×1^ is the SNP effects on the complex trait, and 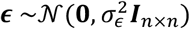 is the environment noise.

In theory, **Δ** can represent any *cis* or *trans* molecular QTLs influencing the perturbed gene, provided that the effect on the gene influences the expression of downstream genes. In this study, we focus on *cis*-eQTLs, as they are the most commonly studied and widely available type of molecular QTLs. Similarly, ***γ*** can represent any gene-to-gene regulatory effect (not only those derived from perturbational screens), as long as it reflects the influence of an upstream gene’s expression on the expression of downstream genes. Here, we focus on effects estimated from CRISPR-based perturbation screens, because these experiments provide causal evidence of regulatory relationships, while traditional association analyses (e.g., steady state co-expression) mainly capture correlation^28^.

Our goal is to test the mediating effect *α* for the focal gene. We obtain the marginal effect size estimate 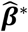 from GWASs, the marginal effect size estimate 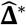 from *cis*-eQTL studies, and perturbation effect size estimate 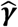 from perturbational screening experiments, respectively. Following previous works^6,29^, we assume 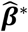 have a sampling distribution 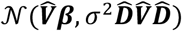 and derive the unbiased and maximum likelihood estimator (MLE) for *α* as

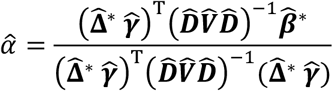

where 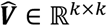 is the estimated SNP correlation matrix (i.e., LD matrix), *σ*^2^ ∈ ℝ_+_ is a heterogeneity parameter accounting for noises due to potential horizontal peliotropy, 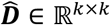 is a diagonal matrix with the standard errors for 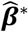 from GWASs. To estimate the standard error for 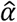, we permute perturbation effects 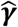 to construct the null distribution for 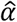, providing a conservative measurement. This null assumes no association between complex traits and mediating genes, meaning any observed relationship is due solely to random perturbation effects rather than regulation by the perturbed genes and their *cis*-eQTLs. We provide detailed descriptions of the model and inference framework in Methods and Supplementary Notes. Mr. PEG is implemented as an open-source command-line Python software available at https://github.com/gusevlab/mrpeg.

### Mr. PEG is well calibrated and well powered in realistic simulations

We first evaluated the performance of Mr. PEG through realistic simulations by varying key parameters, including GWAS sample sizes, *cis*-eQTL sample sizes, the number of *cis*-eQTLs per perturbed gene, the trait heritability explained by a single mediating gene, and the number of perturbed genes. In simulations, we directly used perturbation effect sizes estimated from real data to reflect perturbation effects on downstream focal genes. We compared four versions of Mr. PEG: (1) using MLE with its analytically derived standard error (denoted as “Standard”), (2) using MLE with standard error estimated from permutation tests (“Permutation”), and (3–4) using either an in-sample or an out-of-sample LD matrix to reflect practical scenarios (**Methods**). Under the null scenario, where perturbed genes influence complex traits only through their own expression levels rather than via downstream mediating genes (**Fig. 1**), we observed that Mr. PEG using in-sample LD maintained a calibrated false positive rate of 5% across all parameters (**Figs. 2a, b, Supplementary Fig. 1**). In contrast, the Standard version using out-of-sample LD exhibited moderate inflation in the false positive rate, consistent with previous findings that LD mismatch can inflate association statistics^30^. Notably, the Permutation version effectively corrected for this inflation when using out-of-sample LD, supporting its use as the recommended approach for real data analysis.

**Fig. 2:**
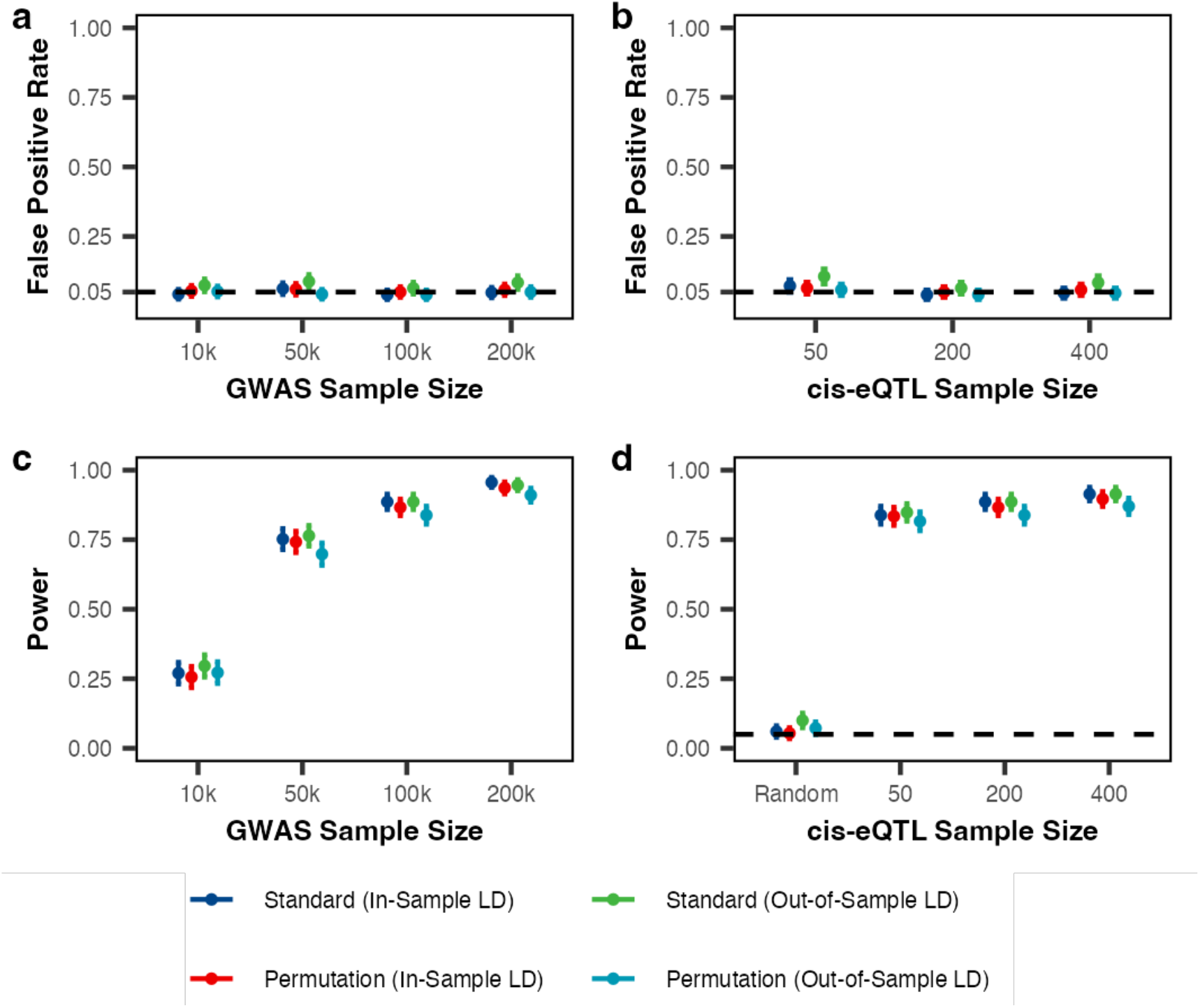
Mr. PEG controls the false positive rate at 5% in the absence of perturbation effects, with power improving with larger GWAS and *cis*-eQTL sample sizes. We compared four versions of Mr. PEG: (1) using standard MLE with its analytically derived standard error (“Standard”), (2) using MLE with standard error estimated from permutation tests (“Permutation”), and (3–4) using either an in-sample or an out-of-sample LD matrix (**Methods**). We simulated scenarios both under the null (no perturbation effects; “Null” in **Fig. 1**) and under the alternative (with perturbation effects; “Alternative” in **Fig. 1**). Each parameter setting was evaluated using 500 independent replicates. The false positive rate was calculated as the proportion of replicates (out of 500) with a P value < 0.05 under the null scenario. Power was calculated as the proportion of replicates (out of 500) with a P value < 0.05 under the alternative scenario. Unless otherwise specified, the default parameters were: GWAS sample size = 100,000; *cis*-eQTL sample size = 200; number of *cis*-eQTLs = 1; number of perturbed genes contributing to the complex trait = 400; number of perturbed genes tested in inference = 400; per-gene trait heritability = 3 × 10^−4^; no pleiotropic effects; and the mediating gene has no *cis*-eQTLs. The points are the mean across simulations, and the error bars are their corresponding 95% confidence intervals. “Random” in plot (d) means the cis-eQTL statistics was replaced by numbers generated from standard normal.

In simulations under the alternative scenario, where perturbed genes influence a mediating gene that influences the complex trait (**Fig. 1**), Mr. PEG achieved high power across a range of parameter settings (**Fig. 2, Supplementary Fig. 2**). As expected, power increased with larger GWAS or *cis*-eQTL sample sizes (**Figure 2c, d**). For a realistic scenario where the GWAS trait exhibited per-gene heritability of 3 × 10^−4^, with a typical GWAS sample size of 200,000 and a *cis*-eQTL sample size of 200, the Permutation version with out-of-sample LD achieved a power of 91.0% (SE=1.3%). In general, increasing the *cis*-eQTL sample size yielded a smaller gain in power compared to increasing the GWAS sample size. For a fixed trait heritability explained by a single mediating gene, increasing the number of perturbed genes reduced power (**Supplementary Fig. 2b**), likely reflecting individually weaker regulatory mechanisms. Finally, to evaluate the calibration when *cis*-eQTL data is missing or erroneous, we replaced the *cis*-eQTL statistics with effects randomly generated from standard normal and observed a well-calibrated null (**Fig. 2d**).

We further evaluated Mr. PEG’s performance under model misspecifications, considering both null and alternative scenarios. First, when pleiotropic effects from *cis*-eQTLs of perturbed genes contributed to trait heritability (**Supplementary Fig. 3a**), power decreased as these effects explained a larger proportion of heritability (**Supplementary Fig. 4a**). For example, average power decreased non-significantly from 83.8% (SE=1.7%) to 77.0% (SE=1.9%) as the proportion explained by pleiotropic effects increased from 0% to 40%. However, this reduction in power became more substantial, from 77.0% (SE=1.9%) to 40.0% (SE=2.1%), as the proportion increased from 40% to 80%. Second, Mr. PEG assumes that mediating gene expression levels are influenced solely by perturbed genes, without contributions from the gene’s own *cis*-eQTLs. To test violations of this assumption, we simulated scenarios in which a mediating gene’s *cis*-eQTL affects its expression and thereby influences downstream traits (**Supplementary Fig. 3b**) and found that power decreased as the proportion of gene expression heritability explained by *cis*-eQTLs increased (**Supplementary Fig. 4b**). However, we found this decrease in average power was not substantial, from 83.8% (SE=1.7%) to 76.8% (SE=1.9%), under a realistic case in which the proportion of gene expression heritability explained by *cis*-eQTLs ranges from 0% to 40%. Both cases highlight that Mr. PEG’s power depends on the proportion of trait heritability explained by perturbation effects. We observed similar patterns when including irrelevant perturbation effects (**Supplementary Fig. 3c, 4c**), or when relevant perturbation effects were absent (**Supplementary Fig. 3d, 4d)**. Importantly, we observed that Standard version with in-sample LD or Permutation version with out-of-sample LD maintained properly calibrated (5.1%; SE=0.2%) false positive rates across all these model misspecification scenarios (**Supplementary Figs. 5**). In addition, we found that model estimates were highly consistent when using either *cis*-eQTL effect sizes or eQTL Z statistics (**Supplementary Fig. 6**). Lastly, we computed the bias of the mediating effect estimator 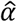, which was 8.78 × 10^−6^ (SE = 1.23 × 10^−5^) across simulations, suggesting that the estimator is approximately unbiased.

Overall, our simulations confirm that Mr. PEG is both well calibrated and highly powerful when model assumptions are met and maintains robustness under varying degrees of model misspecification.

### Mr. PEG identifies mediating genes under evolutionary constraint

We obtained 43 GWAS datasets (average effective sample size: 228,500) on European ancestry (EUR) individuals, including 11 autoimmune diseases, 18 blood-related traits, and other complex diseases (**Supplementary Table 1**). We used *cis*-eQTL data from eQTLGen^27^ Phase I (n=31,684) measured in whole blood from EUR individuals. For perturbation screening data, we obtained gene-to-gene effect size estimates from Yao et al.^25^ where Perturb-seq experiments were performed in macrophage cell lines (MCL), comprising 459 perturbed genes and 13,159 downstream genes, and perturbation effect sizes were inferred using the FR-Perturb method (**Methods**). We confirmed MCL’s relevance to the analyzed complex traits, with 37 of 43 traits showing significant heritability enrichment and the remainder showing nonsignificant enrichment (meta-analyzed enrichment of 1.79 across 43 traits; **Supplementary Fig. 7**). Since the gene-to-gene effect size estimates from perturbation screens can be noisy for smaller effects due to limited power^25,26^, and the true number and identity of perturbed genes for each mediating gene are unknown in practice, we ran Mr. PEG using two types of perturbation effect size matrices: (1) the full matrix, which contains all original values (referred to as the “full matrix”), and (2) a sparse matrix, which retains only the top 1% of absolute effect sizes and replaces the remaining values with 0 (referred to as the “sparse matrix”). A previous study has demonstrated that top absolute effects tend to replicate more consistently across different inference and parameter settings^25^.

Across 43 GWAS, Mr. PEG identified 546 significantly mediating genes (347 unique) after Bonferroni correction for 23,000 tested genes (approximately the number of protein-coding genes in the human genome; **Supplementary Fig. 8**; **Supplementary Table 2**), based on the combined results from both the full and sparse matrices. The sparse matrix identified substantially more significant mediating genes than the full matrix (451 vs. 95), likely indicating that removing estimation noise (i.e., setting small absolute effect sizes to zero) increases statistical power. However, we observed a correlation of 0.32 (P < 1 × 10^−200^) between Mr. PEG effect sizes derived from the full matrix and the sparse matrix, increasing to 0.47 (P < 1 × 10^−200^) when comparing Mr. PEG statistics. Notably, autoimmune diseases exhibited 4.7-fold and 2.8-fold greater absolute effect sizes, on average, compared to blood-related traits and other diseases, respectively (**Supplementary Fig. 9**), likely reflecting a stronger context match between autoimmune diseases and the MCL used in the Perturb-seq experiment.

Mr. PEG model assumes that perturbed genes influence the expression of mediating genes, which in turn affect complex traits (**Fig. 1**). We hypothesize these mediating genes (i.e., Mr. PEG genes) to be functionally closer to the traits studied or under evolutionary constraint. To test these hypotheses, we leveraged multiple metrics that correlate with evolutionary constraint and loss-of-function (LoF) mutation intolerance, including pLI^31^, LOEUF^32^, EDS^33^, s_het_^34^, RVIS^35^, and hs^14,36^, and accounted for confounding factors such as the number of large-effect perturbed genes and the number of significant GWAS signals (**Methods**). We found that Mr. PEG genes were significantly more constrained than the background genes (i.e., 13,159 downstream genes tested; **Fig. 2**; 6 out of 6 tests P < 0.05), with the strongest constraint observed under the EDS metric (P = 2.8 × 10^−10^) which quantifies the overall size, complexity, and redundancy of a gene’s non-coding regulatory architecture^33^. For example, Mr. PEG identified 16 mediating genes associated with rheumatoid arthritis^37^, which showed an average EDS value of 0.63 (SE=0.04) and an average increase of 0.12 more values of EDS (SE=0.02; P = 3.15 × 10^−6^) compared with all other background genes. We further validated these results using a permutation-based approach to mitigate the outlier effects and found similar findings (5 out of 6 tests P < 0.05).

Last, to ensure that the Mr. PEG genes we identified were properly controlled for false positives, we shuffled effect sizes across SNPs from inflammatory bowel disease (IBD) GWAS by de Lange et al.^38^ (GWAS heritability: 0.34), applied Mr. PEG using real *cis*-eQTL and perturbation screening data, and observed well-calibrated results (**Supplementary Fig. 10**).

### Mr. PEG genes exhibit unique molecular features

We compared Mr. PEG-identified genes to those from several alternative methods, including Summary-based Mendelian Randomization (SMR), *trans*-SMR, *trans*-MR, GWAS closest genes, and burden tests (**Supplementary Table 3**; **Methods**). Specifically, SMR identifies genes whose predicted expression is associated with complex traits by leveraging GWAS summary statistics and top *cis*-eQTLs^8^. For *trans*-SMR, we replaced each gene’s top *cis*-eQTL with its top one *trans*-eQTL across all autosomes from eQTLGen. For *trans*-MR, we applied traditional Mendelian Randomization using all significant *trans*-eQTLs for each tested (putatively mediating) gene. We also annotated each downstream gene (i.e., 13,159 downstream genes tested in Mr. PEG) with their top GWAS signals within their 1-M genomic window and curated the closest gene to each GWAS lead variant. Finally, we included gene sets identified through LoF burden tests in GENEBASS (39 out of 43 traits available) to investigate the consistency of effect direction with rare coding variants significantly associated with complex traits (**Data availability**).

Overall, Mr. PEG identified fewer genes than other MR-based methods but more genes than LoF burden, and the identified genes were largely unique (**Supplementary Fig. 11**). For example, SMR and *trans*-MR identified 22.6-fold and 2.6-fold more genes than Mr. PEG, respectively. In addition, 93% of Mr. PEG genes were unique, meaning they were not identified by any other method, whereas this “uniqueness” across other methods was 71% on average. Previous studies have shown that MR-based methods can produce false positives due to confounding factors such as *cis*-eQTL linkage disequilibrium and pleiotropy^39^. We hypothesize that *trans* effects estimated through CRISPR-based perturbation screens may be less susceptible to this tagging issue, resulting in fewer significant genes. Conceptually, *trans*-MR is most similar to Mr. PEG, as both leverage whole-genome *trans* effects to identify genes associated with complex traits, with *trans*-MR based on *trans*-eQTL data and Mr. PEG on perturbational data. However, we observed generally low and non-significant correlations between *trans*-MR and Mr. PEG statistics (**Supplementary Table 4**), suggesting that perturbational screening data may capture regulatory mechanisms distinct from those identified by *trans*-eQTL analyses, consistent with previous findings^25^.

To investigate the functional features of genes identified by these methods, we analyzed gene annotations curated by Mostafavi et al.^14^, and compared them using significant genes identified by each method (**Methods**). First, we found that Mr. PEG genes exhibited longer average enhancer lengths across active biosamples than genes identified by other methods, based on enhancer measurements from Nasser et al.^40^ and the ROADMAP project^41^ (**Fig. 4a, Supplementary Figure 12a**). This suggests that Mr. PEG genes may harbor more transcription factor (TF) binding sites and therefore possess a more complex regulatory architecture, capable of receiving signals from multiple pathways. Second, we observed that Mr. PEG genes are more likely to be co-expressed within gene networks compared to genes from other methods, except for *trans*-MR, which showed even stronger co-expression patterns (**Fig. 4b**). This is consistent with the previous findings that top co-expressed gene pairs are enriched in *cis*-by-*trans*-eQTLs^25^. Third, Mr. PEG genes appear to be more constrained than those identified by most other methods (mean=0.08; se=0.005), except for genes nearest to GWAS loci (mean=0.09; se=0.001), based on the *hs* annotation^14^, which directly quantifies selection (**Fig. 4c**)^36^. Notably, genes identified by *trans*-MR, the method most similar to Mr. PEG, exhibited the lowest *hs* values and were depleted relative to background (mean=0.05; SE=0.003), suggesting that *trans*-eQTL–informed genes are less intolerant to LoF mutations, consistent with previous findings^14^. We observed similar trends using other constraint-related metrics such as pLI, s_het_, EDS, LOEUF, and RVIS (**Supplementary Figs. 12b-h**). Overall, these patterns likely reflect the different regulatory roles of genes within molecular pathways, as identified by each method.

**Fig. 3:**
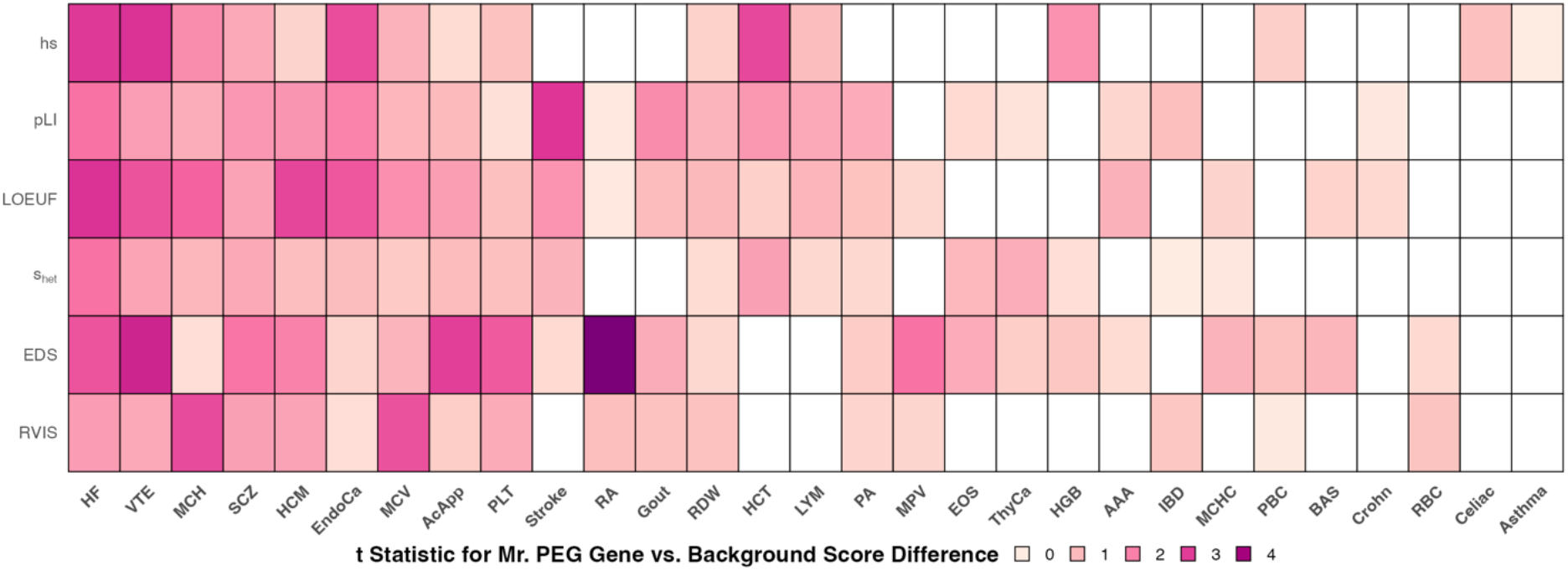
Mr. PEG genes exhibit greater constraint compared to background genes. We include only traits (x-axis) with more than three Mr. PEG–significant genes. For each metric (y-axis), we performed a linear regression in which the dependent variable is the gene’s metric score and the independent variable is a binary indicator for whether the gene is Mr. PEG–significant, adjusted for our three covariates: the indicator for using the full effect-size matrix versus the sparse matrix, the number of non-zero perturbation effects, and the number of significant GWAS variants. The color indicates the magnitude of the t-statistic, with negative t-scores shown in white. LOEUF and RVIS statistics are flipped to align their directionality with the other four metrics that correlate positively with gene constraint.

**Fig. 4:**
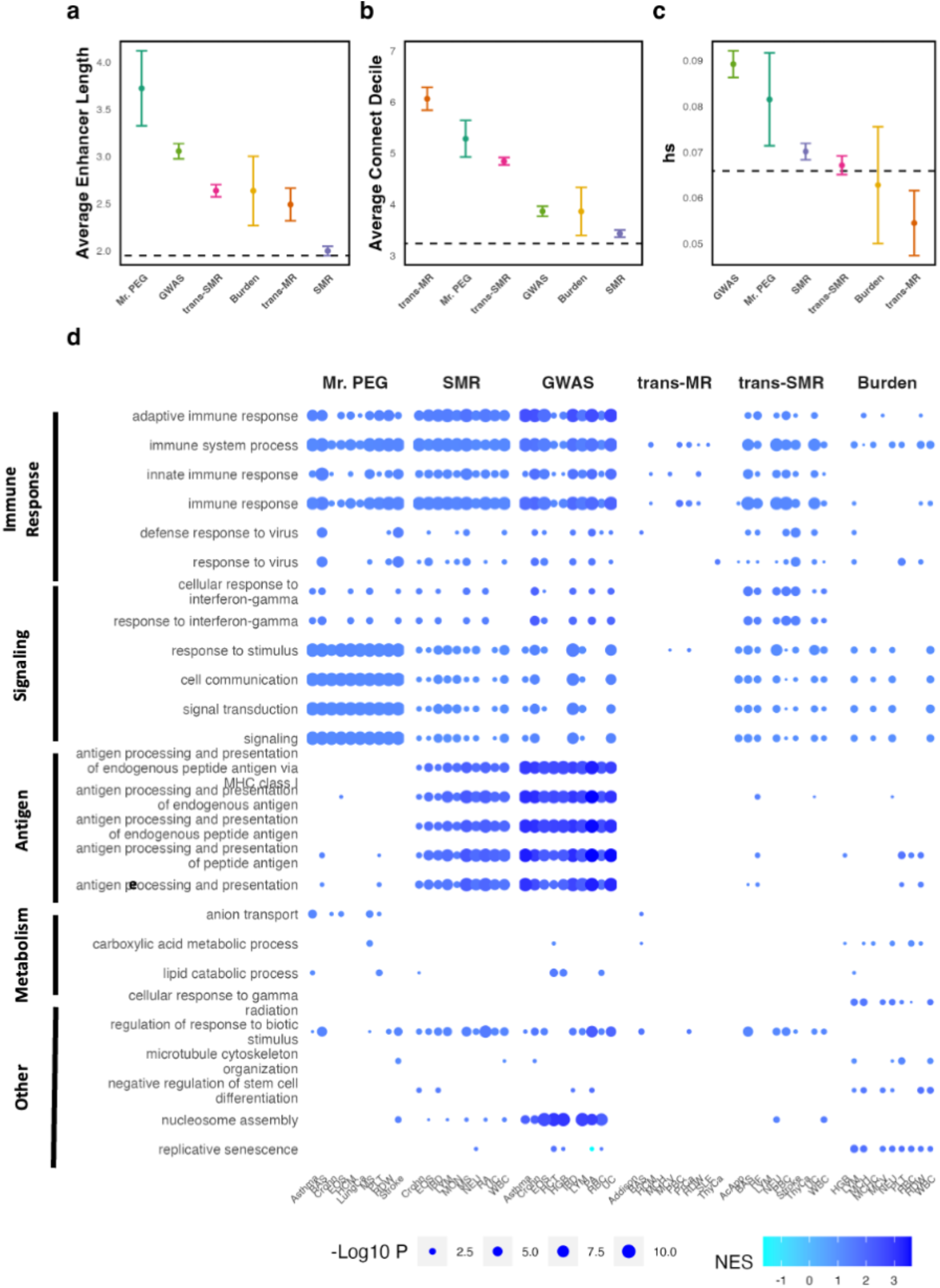
Mr. PEG exhibits different molecular features from GWAS, MR-based, and burden tests. (a) Average enhancer length annotation prepared by Mostafavi et al. using data from the ROADMAP project. (b) Connect decile annotation prepared by Mostafavi et al., where higher values indicate that a gene co-expresses with more genes. (c) hs annotation prepared by Mostafavi et al., where higher values indicate stronger selection. (d) The top five most enriched Gene Ontology Biological Process terms across all complex traits (y-axis), and the top five most enriched traits across all terms for each method (x-axis). NES stands for normalized enrichment scores. Non-significant terms are shown in white. P value is not adjusted and two sided computed from GSEA.

We further extended our enrichment analysis to include all genes by treating the absolute value of the association statistic of each method as continuous annotation. This approach revealed clear distinctions among the genes ranked by different methods, as shown by Gene Set Enrichment Analysis (GSEA) using Gene Ontology (GO) Biological Process terms^42,43^. First, Mr. PEG genes were enriched for 8,387 significant terms (FDR-adjusted P < 0.05), representing a 1.7-fold increase over SMR, 1.2-fold over gene annotations based on the most significant GWAS signals, 16-fold over *trans*-SMR, 559-fold over trans-MR, and 12.1-fold over the LoF burden test (**Data Availability**). Second, genes annotated by Mr. PEG statistics were predominantly enriched for immune response and signaling-related terms (**Fig. 4d**; **Data Availability**). In contrast, genes annotated by GWAS and SMR statistics were mainly enriched for immune response and antigen-related terms but showed weaker enrichment for signaling-related terms compared to Mr. PEG. These results suggest that Mr. PEG, by leveraging perturbational screening data, may capture genes involved in complex trait pathways that are often overlooked by traditional GWAS and MR-based methods.

The sign of the Mr. PEG statistic reflects whether upregulation (negative sign) or downregulation (positive sign) is associated with higher levels or increased risk of a complex trait. To examine differences between upregulated and downregulated genes identified by Mr. PEG, we ranked genes by their nominal statistics and repeated the GSEA analysis. For immune related diseases, genes where higher expression was linked to increased disease risk were mainly enriched in transcriptional regulatory processes, whereas genes whose lower expression increased disease risk were enriched in nucleotide metabolic processes and energy production (**Supplementary Fig. 13**). This may suggest that reduced nucleotide and energy metabolism may limit the capacity of immune cells to expand and function effectively. In contrast, for blood-related and other complex traits, genes with lower expression linked to elevated trait levels and increased risk were largely enriched in transcriptional regulatory processes and nucleic acid metabolic processes, while genes with higher expression linked to elevated trait levels or disease risk were enriched in immune and defense activation pathways (**Supplementary Fig. 14-15**). Overall, these patterns suggest regulatory heterogeneity across trait categories. In certain biological contexts, increased expression of regulatory genes may drive immune activation, whereas blood-related traits may depend more on maintaining stable baseline transcriptional programs.

Mr. PEG integrates GWAS and *cis*-eQTL data, in which both are typically based on common variants at the SNP level, together with gene-level perturbational screening data. We hypothesized that this integration could help identify genes whose rare coding variants are associated with complex traits. To test this, we correlated the Mr. PEG effect sizes derived from the sparse Perturb-seq matrix with LoF burden effect sizes across genes and observed a positive association (P = 1.6 × 10^−4^; **Fig. 5a**). In contrast, we did not detect significant associations when using SMR effect sizes (P = 0.13), or trans-MR effect sizes (P = 0.46; **Methods**). This result likely reflects that perturbational screens can capture mechanisms that are relevant to both rare and common variants. We caution that when using Mr. PEG Z statistics, where the standard errors were estimated from permutations, instead of using effect sizes directly, the association became less significant (P = 0.04), likely because effect sizes better preserved directional concordance with LoF burden signals. We also observed reduced signal (P = 0.09) when substituting LoF burden effect sizes with their corresponding Z statistics, potentially due to the burden test power driven by gene coding sequence length^21^. To ensure that our associations were not confounded by gene length or coding sequence length, we re-evaluated the analysis after adjusting for each factor separately, and the results remained consistent (P = 8.7 × 10^−4^ and 1.4 × 10^−4^, respectively). In contrast, using the full Perturb-seq effect size matrix did not yield a significant association with LoF burden effect sizes (P = 0.75), likely due to reduced statistical power.

**Fig. 5:**
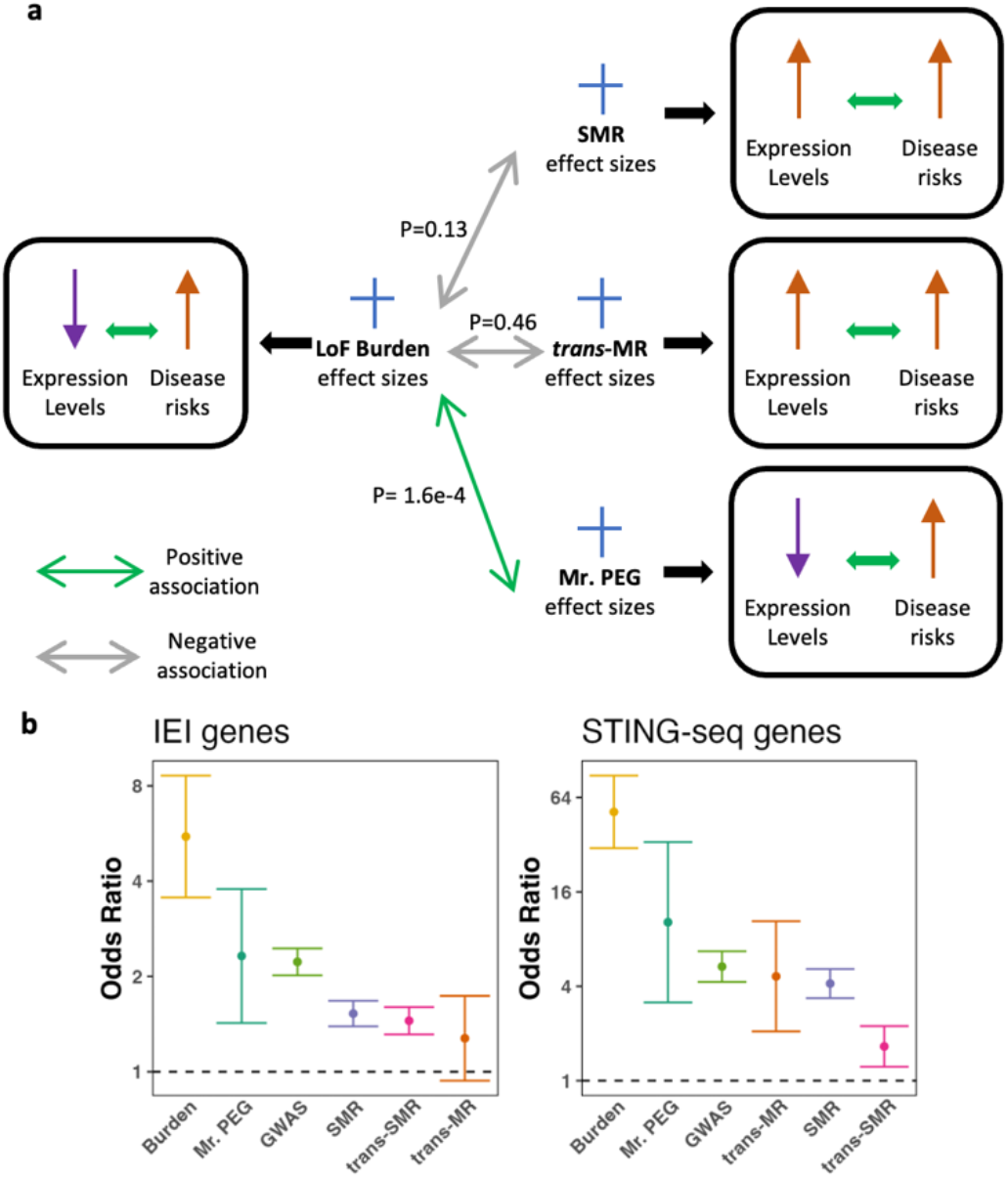
Mr. PEG statistics are positively associated with loss-of-function burden effect sizes. (a) Positive loss-of-function (LoF) effect sizes indicate that decreased gene expression is associated with increased disease risk. Similarly, positive Mr. PEG effect sizes reflects the same direction of effect. In contrast, positive SMR and trans-MR effect sizes indicate that increased gene expression is associated with increased disease risk. P-values are calculated using linear regression adjusted for traits and are two-sided. (b) Logistic regression was performed separately for each method, where the dependent variable indicates whether a gene is an inborn errors of immunity (IEI) or STING-seq gene, and the independent variable indicates whether the gene is identified as significant by that method. Results were meta-analyzed across all traits for IEI genes and across all blood-related traits for STING-seq genes, respectively. Points represent the meta-analyzed enrichment estimates, and error bars denote the corresponding 95% confidence intervals.

**Fig. 6:**
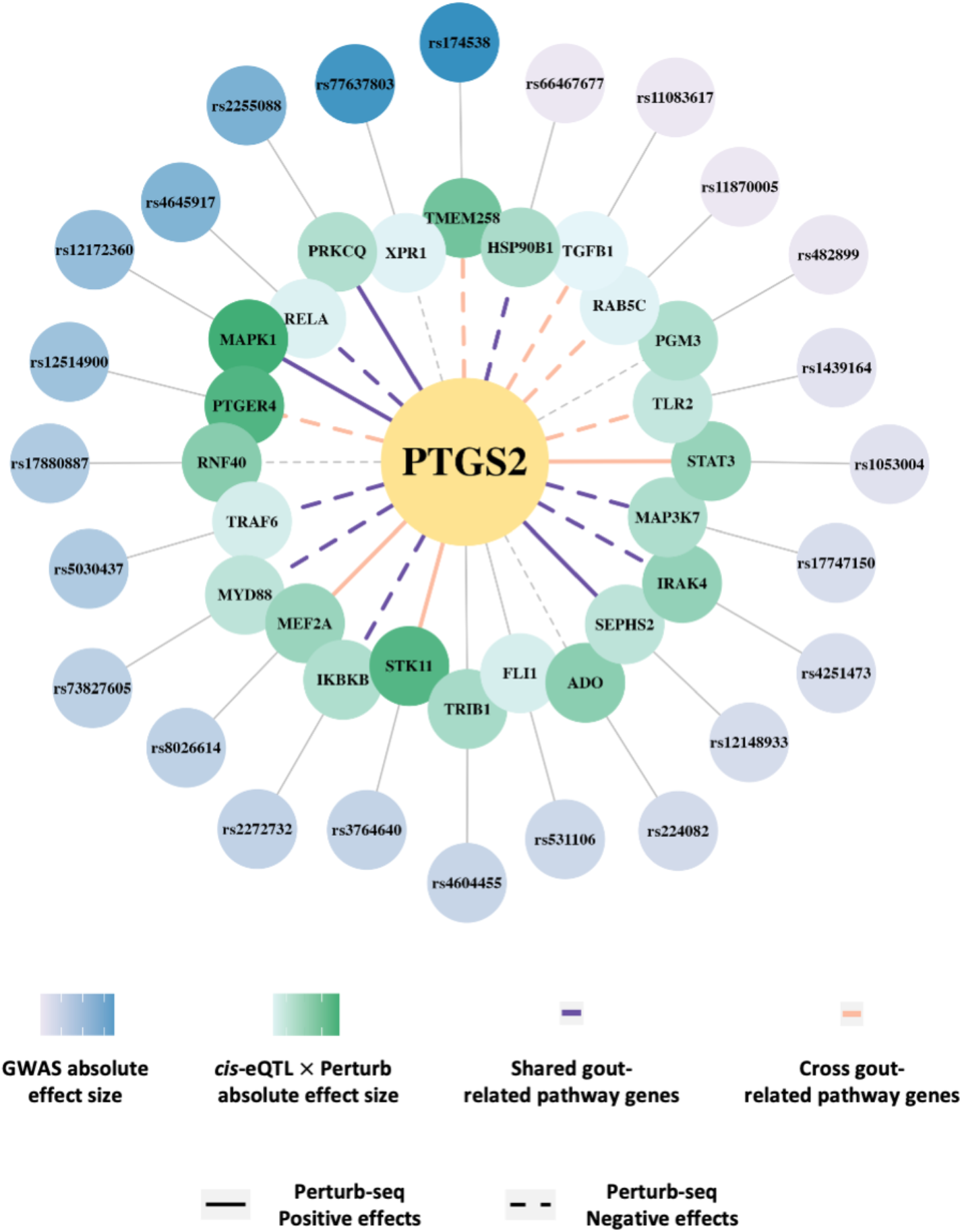
Mr. PEG identifies the *PTGS2* gene as a putative mediating gene for gout. Perturbed genes of *PTGS2* are shown in green, with shading indicating the product of *cis*-eQTL effect size and Perturb-seq effect size. Perturbed genes are defined as those with effect sizes in the top 1% of the original Perturb-seq matrix. The top *cis*-eQTL for each perturbed gene (i.e., the most significant) is shown in blue, with shading representing the absolute GWAS effect size. Genes are ordered by the GWAS effect size of their top *cis*-eQTL. Colored lines (purple or orange) connect genes appearing in KEGG pathways related to gout: purple indicates both genes are in the same pathway, while orange indicates they are in different but closely associated pathways. Solid lines represent positive Perturb-seq effects; dashed lines indicate negative effects. None of the genes are SMR significant, and none of the *cis*-eQTLs are GWAS significant.

Last, we used two curated gene sets to further validate the association between Mr. PEG genes and LoF burden effects. We first used Inborn Errors of Immunity (IEI) genes, in which rare germline mutations result in immune system dysfunction^44^. Mr. PEG genes showed significant enrichment for IEI genes across all complex traits (meta-analyzed OR = 2.32; P = 7.12e-4). Mr. PEG genes were more enriched, although not significantly, relative to all other methods except for the LoF Burden genes (**Fig. 5b**). The relatively larger standard error is likely attributable to the smaller number of Mr. PEG genes compared with other methods. Second, we used *cis*-targeted genes for blood traits identified by STING-seq^45^, a framework that integrates GWAS data, CRISPR screens, and single-cell sequencing. This gene set provides high-confidence causal genes for blood-related traits, and we found similar results: Mr. PEG genes for blood-related traits showed greater enrichment for STING-seq–identified gene sets than all methods except Burden genes (**Fig. 5b**), although the difference was not statistically significant due to the limited number of Mr. PEG genes.

Together, these findings demonstrate that Mr. PEG-based effect sizes, inferred from common variant GWAS data and integrated with perturbational screening data, correlated with rare coding burden effects and capture complementary mechanisms not detected by *cis*- and *trans*-eQTL-based approaches.

### Mr. PEG identifies mediating genes with drug repurposing potential

Lastly, we highlight three examples of Mr. PEG genes that consistently exhibit “mediating gene”-like features (**Fig. 5**). First, Mr. PEG identified *PTGS2* as a putative mediating gene for gout (P = 1.02 × 10^−8^). *PTGS2* (GRCh38: chr1:186,671,791–186,680,922) encodes the COX-2 enzyme, a key regulator of inflammatory responses. Gout, a form of inflammatory arthritis, is characterized by sudden joint pain and swelling.

We validated the association through multiple lines of evidence. First, several independent studies have reported that the upregulation of *PTGS2* plays a significant role in inflammation associated with gout,^46–49^ consistent with our findings by Mr. PEG (effect size=−1.9 × 10^−3^; a negative sign indicates that upregulation associated with disease risks). In addition, we found that several medications relating *PTGS2* are effective against gout. For instance, celecoxib, a COX-2 inhibitor, has demonstrated therapeutic benefit^50^. *PTGS2* is also the molecular target of N02BA03-class anti-inflammatory drugs, such as choline salicylate, which inhibits COX-2 and may likewise contribute to gout management^51^. To support this, we conducted generalized summary-data-based Mendelian randomization (GSMR) analysis^52,53^ between N02BA drug usage^54^ and gout. We found a significant negative association (effect size=-0.23; P = 3.0 × 10^−3^), indicating that higher genetic liability to N02BA drug usage is associated with a reduced risk of gout. This finding may suggest that *PTGS2* can serve as a genetic mediator linking N02BA03-class drug actions to the therapeutic or protective effects against gout.

Second, in the GWAS dataset^55^ used by Mr. PEG to identify *PTGS2*, the most significant SNP had a P value of P = 7.0 × 10^−3^. However, in a later and larger meta-analysis of GWAS across multiple ancestries (sample size increase of 62,475; **Data Availability**), the P value of the most significant SNP improved to P = 7.0 × 10^−6^, suggesting that the gene may be a genuine association but threshold-dependent. Third, we found that *PTGS2* is highly intolerant to loss-of-function mutations, as indicated by a pLI score of 1 (**Data Availability**), and is under strong selective constraint, ranking in the top 25% of *hs* values across all genes.

Next, *PTGS2* is involved in several KEGG pathways relevant to gout such as TNF-α signaling pathway, IL-17 signaling pathway, NF-κB signaling pathway. For example, it functions as a downstream target in the NF-κB signaling pathway, a key regulator of the inflammatory response in gout. Upstream genes of *PTGS2*, including *RELA, PRKCQ, TRAF6, MYD88, IKBKB, IRAK4*, and *MAP3K7*, are all components of the NF-κB pathway (hsa04064) and are utilized in Mr. PEG’s inference framework to identify *PTGS2* as a mediating gene (**Fig. 5**; **Supplementary Table 5, 6**). Lastly, we confirmed that *PTGS2* was not significant in *trans*-MR, SMR, *trans*-SMR, or LoF burden analyses, underscoring Mr. PEG’s ability to identify distinct mediating genes for complex diseases.

As another example, Mr. PEG uniquely identified *TNFAIP3* (also known as *A20*; GrCh38: chr6:137,866,349-137,883,314; **Supplementary Figure 16**) as a putative mediating gene for heart failure (P= P = 4.4 × 10^−8^). Inflammatory signals, particularly those involving the TNF-α and NF-κB pathways, can upregulate *TNFAIP3* to shut down the previous pathways in negative feedback loop^56^. This aligns with our finding in Mr. PEG that the upregulation of *TNFAIP3* was associated with heart failure risks (effect size= −1.4 × 10^−3^). In addition, L04AB drugs (TNF-α inhibitors) typically also function as immunosuppressants, by blocking TNF-α signaling. Using GSMR analysis, we found that higher genetic liability to L04AB drug usage was associated with an increased risk of heart failure (P = 1.2 × 10^−7^). This observation may suggest a complementary role between *TNFAIP3* and L04AB drugs in controlling inflammation, where dysfunctional *TNFAIP3* could increase inflammation and heart failure risk, and TNF-α inhibitors may help compensate by suppressing inflammatory activity. Furthermore, similar to the *PTGS2* case, the significance of the top SNPs within the *TNFAIP3* locus increased when comparing the GWAS used for inference to a subsequent, larger GWAS that included 99,838 additional samples (P = 0.2 vs. 1.2 × 10^−4^). *TNFAIP3* is also highly constrained, as indicated by a pLI score of 1. Additionally, several upstream perturbed genes of *TNFAIP3* are involved in KEGG pathways known to play a role in heart failure (**Supplementary Table 5, 6**), such as the NF-κB signaling pathway (hsa04064) and NOD-like receptor signaling pathway (hsa04621).

Lastly, Mr. PEG uniquely identified *ATP1B1* as a putative mediating gene for hypertrophic cardiomyopathy (HCM; GrCh38:chr1:169,105,697-169,310,992; **Supplementary Figure 17**). HCM is a genetic disease characterized by abnormal thickening of the heart muscle, most caused by mutations in key genes such as *MYH7* and *MYBPC3*. Although a direct association between HCM and *ATP1B1* has not been established through experiments, we suspected that the downregulation of *ATP1B1* might worsen the development of HCM based on Mr. PEG’s effect size of 6.25e3 (P = 2.1 × 10^−6^). *ATP1B1* encodes the β-1 subunit of the membrane ion pump responsible for sodium–potassium transport across the cell membrane. Reduced *ATP1B1* expression can potentially disrupt intracellular sodium balance, leading to sodium accumulation that may exacerbate cardiomyopathy. Furthermore, C03-class drugs inhibit sodium reabsorption in renal tubules to lower blood pressure and can serve as supportive therapy to help relieve symptoms in cardiomyopathy. Using GSMR analysis, we found that higher genetic liability to C03 drug use is associated with higher risk of HCM (P = 0.02). Similar to the *TNFAIP3* example, this may suggest a complementary relationship between *ATP1B1* and C03 drugs: *ATP1B1* dysfunction promotes sodium retention, worsening HCM, while C03 drugs may help ease this effect. In addition, the significance of the top SNPs within the *ATP1B1* locus increased when comparing the GWAS used for inference to a later, larger GWAS with 37,692 additional samples (P = 0.29vs. P = 3.3 × 10^−4^). This gene is also highly constrained, as indicated by a pLI value of 1. Importantly, *ATP1B1* and its upstream perturbed genes are involved in several KEGG pathways potentially associated with HCM (**Supplementary Table 5, 6)**, including adrenergic signaling in cardiomyocytes (hsa04261), cardiac muscle contraction (hsa04260), cAMP signaling pathway (hsa04024), and cGMP PKG signaling pathway (hsa04022).

These results suggest that Mr. PEG leverages perturbational screening data to identify biologically meaningful mediating genes not detected by GWAS, MR-based, or burden tests, highlighting its potential for informing drug repurposing in disease treatment.

## Discussion

In this study, we present Mr. PEG, a novel yet straightforward framework that integrates perturbational screens, *cis*-eQTL, and GWAS summary data to identify mediating genes for complex traits. Mr. PEG models the complex trait as a function of the expression levels of mediating genes, which in turn are modeled as a linear combination of perturbed gene expression levels. Through extensive simulations under realistic conditions, we demonstrated that Mr. PEG is well calibrated under the null scenarios, achieving 5% false positive rates when no perturbation effects are present, and shows 91% (SE=3%) power when upstream perturbations influence mediating genes with 200,000 GWAS sample sizes.

We applied Mr. PEG to a broad set of GWAS datasets and identified 546 mediating genes across 40 complex traits. These genes are more constrained than background genes, likely suggesting closer functional proximity to complex traits. Compared to genes identified by most GWAS and MR-based methods, Mr. PEG-prioritized genes tend to exhibit longer average enhancer lengths across active biosamples and stronger co-expression patterns within networks. Gene set enrichment analysis reveals that Mr. PEG identifies a strikingly higher number of enriched GO terms compared to MR-based methods burden tests, suggesting that Mr. PEG, by leveraging perturbational screening data, captures genes involved in complex trait pathways missed by traditional methods. Notably, Mr. PEG effect sizes learned from common non-coding variant GWAS correlate with rare coding burden effects, unlike other methods, highlighting its ability to capture complementary mechanisms missed by *cis*-eQTL-informed methods. As an illustrative example, Mr. PEG identified *PTGS2* as a putative mediating gene for gout, which was not detected by other methods.

We acknowledge several caveats and limitations to our real data analysis. First, while the Mr. PEG model may appear conceptually similar to the omnigenic model proposed by Boyle et al.^18^ and Liu et al.^17^, it also has some key differences. The omnigenic model assumes that a set of core genes influences complex traits through direct effects, while many peripheral genes affect the trait indirectly by regulating those core genes. In contrast, Mr. PEG does not assume that the mediating genes it identifies are core genes, nor does it treat perturbed genes as peripheral by default. In fact, the genes knocked out in our Perturb-seq dataset (i.e., Yao et al.^25^) were manually selected as important immune-related genes and may be more consistent with the role of core genes under the omnigenic assumptions. Mr. PEG aims to identify genes whose expression levels are associated with complex traits and are influenced by upstream regulators, as captured through CRISPR experiments, without assigning any predefined importance or hierarchy. Therefore, while classically core genes are expected to show significant Mr. PEG effects, Mr. PEG will also identify other types of genes that mediate between (or are consistently pleiotropic with) *cis*-eQTL genes and the disease.

Second, the Perturb-seq^25^ and *cis*-eQTL^27^ summary data we used may not fully match the biological context of the complex traits being analyzed, which can limit our ability to identify mediating genes. The Perturb-seq data were generated from macrophage cell lines, which are most relevant to immune and inflammation-related conditions, while the *cis*-eQTL data were obtained from whole blood. When the context of these datasets does not align closely with the underlying biology of the analyzed traits, our ability to detect true mediating genes may be reduced. Nevertheless, our current analysis still identified biologically meaningful candidates, such as *PTGS2* for gout. We anticipate that Mr. PEG will become even more powerful as additional context-matched datasets become available in the future.

Third, current perturbation screening datasets vary widely and often do not replicate most effects due to limitations in experimental design, differences in targeted knockouts, and other confounding factors. For example, we find a low correlation (r=0.05) of Perturb-seq effect sizes between the Yao et al.^25^ and Replogle et al.^24^ datasets. This leads to a similarly low concordance of Mr. PEG Z statistics between them, with a correlation of r=0.006 (P = 7.1 × 10^−4^), when using the full Perturb-seq effect size matrix. Users should carefully consider which perturbational screening data are most appropriate for their specific research questions. To support this decision, we provide the effect size matrix used in our analysis as part of the Mr. PEG software package.

Fourth, in our real data analysis, we heuristically use two types of Perturb-seq effect size matrices: the full matrix, which contains all original estimated values, and the sparse matrix, which retains only the top 1 percent of absolute effect sizes. This approach is motivated by two considerations. First, perturbation effect size estimates for small signals are often noisy^25,26^. Second, for a mediating gene given a complex trait, its true number of upstream perturbed genes remains unknown. Our simulation results show that misspecifying the number of perturbed genes, whether specifying more or fewer than the true number, reduces statistical power (**Supplementary Fig. 4**). Our real data analysis shows that the sparse matrix identifies more significantly mediating genes and exhibits stronger correlation with LoF burden effect sizes. Given current limitations in effect size estimation methods for perturbational screens, we recommend that users explore different sparsity thresholds to reduce noise and improve robustness. Alternatively, users may apply shrinkage techniques to the effect sizes to reduce estimated noise.

Lastly, Mr. PEG does not account for directional pleiotropy, where SNPs influence complex traits through pathways that are not mediated by perturbation effects. In the traditional Mendelian randomization framework, methods such as MR-Egger have been proposed to test for directional pleiotropy^57^. However, in practice, detecting such effects is challenging because individual SNPs typically have small effect sizes on complex traits, and the pleiotropic component can be difficult to distinguish, especially when perturbation effects explain a large proportion of the trait variance. In our simulation framework, we observed only a minimal reduction in the power of Mr. PEG as the proportion of variance explained by directional pleiotropy increased from 0% to 40% (i.e., “uncorrelated pleiotropy” between SNPs and mediating effects). However, when correlated pleiotropy exists between SNPs and mediating effects, we instead expect potential false positives^58^. As a result, we acknowledge this as a limitation of the current method and highlight it as an important direction for future work aimed at improving the identification of mediating genes while accounting for both correlated and uncorrelated pleiotropic effects.

Overall, Mr. PEG integrates perturbational screens, *cis*-eQTL, and GWAS data to identify mediating genes for complex traits. We anticipate that it will become an increasingly valuable tool in the genetics field as more perturbation-based datasets become available.

## Methods

### The mediating gene model

We assume that the expression level of a focal gene has a mediating effect *α* ∈ ℝ on the complex trait, and we refer to such a gene as a mediating gene. We further assume that the expression level of the mediating gene can be modeled as a linear combination of the expression levels of a set of upstream genes, which we refer to as perturbed genes. These perturbed genes regulate the mediating gene through the gene regulatory network, and this regulatory effect can be quantified using perturbational screen experiments. Finally, we assume that the expression levels of the perturbed genes are influenced by their corresponding eQTLs. Based on these assumptions, we introduce a Mendelian randomization-like framework that integrates perturbational screens, eQTL data, and GWAS summary data to identify mediating genes for complex traits.

We model a normalized complex trait ***y*** ∈ ℝ^*n*×1^ measured in *n* individuals as a function of the expression levels of a focal gene ***g*** ∈ ℝ^*n*×1^ as

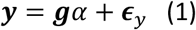

where *α* ∈ ℝ is the mediating effect and 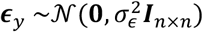 is the environmental noise. We further model the mediating gene expression level ***g*** as a linear combination of *t* perturbed gene expression level matrix ***M*** ∈ ℝ^*n*×*t*^ with a gene-to-gene effect size vector ***γ*** ∈ ℝ^*t*×1^ as

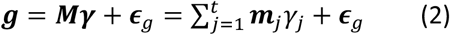

where *j* is the perturbed gene index, ***m***_*j*_ ∈ ℝ^*n*×1^ is the expression level vector of a perturbed gene, *γ*_*j*_ ∈ ℝ is the effect scaler of a perturbed gene on the focal gene, and 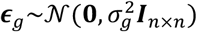 is the environmental noise. Last, we assume the expression levels ***m***_*j*_ is a linear combination of the genotypes (standardized matrix 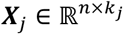) as

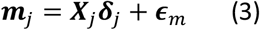

where 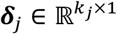 is the eQTL effects, *k*_*j*_ is the number of SNPs for the perturbed gene *j* and 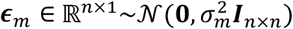 is the environmental noise.

The algebraic expansion (**Supplementary Note**) after plugging (2) and (3) into (1) gives us

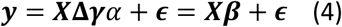

where ***X*** ∈ ℝ^*n*×*k*^ is the genotype matrix across *t* perturbed genes, *k* is the number of unique SNPs in ***X*, Δ** ∈ ℝ^*k*×*t*^is the eQTL effect size matrix of *k* SNPs to *t* perturbed genes, ***β*** ∈ ℝ^*k*×1^ is the SNP effects on the complex trait, and 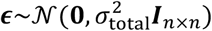 is the aggregated environment noises from (1), (2), and (3). We note that **Δ** is sparse because each perturbed gene (i.e., a column in **Δ**) only has a few eQTLs (i.e., non-zero entries), thus the rest of the entries in the column are zero. If *k* = *t*, each perturbed gene only harbors one eQTL, so **Δ** is reduced to a diagonal matrix.

We also note that **Δ** can be any *cis* or *trans* molecular QTL if the molecular feature of a gene influences the expression of downstream genes. We use *cis*-eQTLs, as they are the most commonly studied type of molecular QTL. Similarly, ***γ*** can be any gene-to-gene regulatory effect (not necessarily those derived from perturbational experiments) if it represents how an upstream gene’s expression affects the expression of downstream genes. Here, we use effects estimated from CRISPR-based perturbational screens, because these experiments provide causal evidence of regulatory relationships, rather than correlation.

### Inference using maximum likelihood estimation

Let 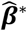 be the marginal effect size estimate for each SNP in GWAS and we assume 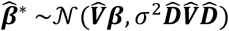 where 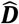 is a diagonal matrix where the entries are the standard error of 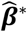 for each SNP, 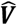 is the estimated SNP correlation matrix (i.e., LD matrix; usual), and *σ*^2^ ∈ ℝ_+_ is the heterogeneity parameter. Based on (4), we have

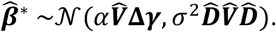

We assume 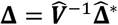 in a well-powered *cis*-eQTL study (e.g., eQTLGen) where 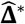 is the marginal *cis*-eQTL effect size estimates. 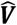 is typically obtained from external reference genotype data. We heuristically assume 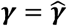 in well-designed experiments, where 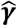 is the gene-to-gene perturbational effects estimated in perturbational screening experiments (see the last paragraph of this section for caveats). We note that the uncertainty of the estimates 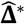 and 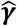 is not modeled. For 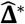, we can instead use the corresponding Z statistics to incorporate uncertainty, and this is less of an issue in a well-powered *cis*-eQTL study. For 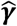, many methods rely on permutation tests to obtain P values because perturbational effect estimates do not follow a normal distribution^25^. Therefore, Z statistics derived from discrete P values in permutation test may not perform well in this setting.

Then, we can derive the maximum likelihood and unbiased estimators for *α* and the unbiased estimator for *σ*^2^ as follows:

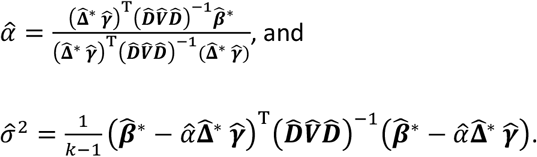

The test statistic for 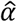 can be derived as:

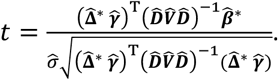

The test statistic follows a *t* distribution with *k* − 1 degrees of freedom.

Alternatively, we can perform a permutation test to empirically construct the null distribution for 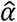 and compute its mean 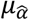 and standard deviation 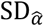. In detail, we shuffle only the rows in the perturbational effect estimates 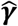 while keeping the SNP matching across *cis*-eQTL data, GWAS data, and the correlation matrix. The constructed null distribution assumes no association between complex traits and mediating genes, meaning any observed relationship is due solely to random perturbation effects rather than regulation by the perturbed genes and their *cis*-eQTLs. Last, when the number of perturbed genes in the model is large (e.g., *k* > 100), we can compute the permutation *Z* statistic as 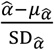, from which a two-sided P value based on the standard normal distribution can be obtained. Alternatively, when *k* is small, the distribution of 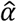 may deviate from normality; as a result, a discrete permutation-based *P* value is preferred instead of computing a *Z* statistic.

To ensure fast and stable inference, we further simplify our model: since the most significant *cis*-eQTL (i.e., top *cis*-eQTL) of a gene typically explains most of its expression signals^7^, we selected only the top *cis*-eQTL for each perturbed gene. As a result, the *cis*-eQTL effect size matrix 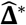 is reduced to a diagonal matrix with top *cis*-eQTL effect sizes with a dimension of *k* × *k* or *t* × *t* as *k* = *t*, so 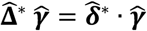 where · means element-wise multiplication. A detailed description of the inference procedure is provided in the **Supplementary Note**.

In practice, 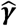 can be obtained in two ways regardless of the effect size estimation algorithm used. One approach is to use marginal estimates, which do not account for across-gene correlation and are, for example, obtained separately for each gene. The other approach is to use conditional estimates, which account for across-gene correlation and are, for example, obtained from a joint model that includes all genes. Ideally, conditional estimates of ***γ*** are preferred because they better reflect gene-to-gene effects in regulatory networks. For example, suppose gene A mediates gene B, and gene B in turn mediates gene C, with gene A affecting gene C only through gene B, and the effect from A to B being small. Under marginal estimation, the effect estimation from A to C may appear larger than the effect from B to C. In contrast, under conditional estimation, the effect estimation from A to C is expected to be smaller than the effect from B to C. In our analysis, we used estimates from Yao et al., who applied FR-Perturb, a factor-analysis-based algorithm incorporating LASSO. These estimates can be viewed as conditional estimates because the method jointly models all perturbed genes through a regularized framework, although the extent to which conditional effects are accurate also depends on the number of perturbations per cell.

### Simulating *cis*-eQTL summary data

Following our previous work^59–61^, we conducted simulations using genotypes from 490 individuals of European ancestry obtained from the 1000 Genomes Project Phase 3 (1000G). For each gene on the 22 autosomes, we extracted the top three significant *cis*-eQTL rsIDs from eQTLGen, ensuring that these variants were also present in the 1000G reference genotypes. To maintain quality, we restricted our selection to high-quality HapMap SNPs, defined as having <1% missingness, minor allele frequency (MAF) >1%, and no significant deviation from Hardy-Weinberg equilibrium (HWE mid-p < 1 × 10^−5^). We then randomly selected a total of *t* ∈ {200, 400, 600,800} genes as perturbed genes across the 22 autosomes. The number of genes selected per autosome was proportional to the distribution of protein-coding genes across the autosomes in the human genome.

For a perturbed gene *j*, to simulate its *cis*-eQTL summary data, we need to generate *cis*-eQTL effect sizes, genotypes, and gene expression levels. First, we simulated *cis*-eQTL effect sizes 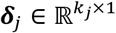 from 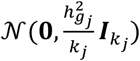 where *k* ∈ {1, 2, 3} is the number of *cis*-eQTLs per perturbed gene, 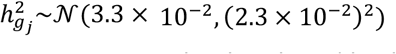 is the expression heritability (i.e., the proportion of variation of the expression level explained by the genotypes for a single perturbed gene), and 3.3 × 10^−2^ and 2.3 × 10^−2^ are the mean and standard error of for 12,160 genes measured in whole blood of Genotype-Tissue Expression study (GTEx) reported by Wheeler et al.^62^ We selected rsIDs from the top three *cis*-eQTLs prepared above in a descending order based on their significance.

To simulate genotypes, we first constructed the LD matrix 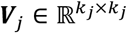 using the selected rsIDs’ genotypes in the reference data, and simulated a centered and standardized genotype matrix 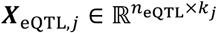 from a multinormal distribution 𝒩(**0, *V***_*j*_) where *n*_eQTL_ ∈ {50, 200, 400}. Next, we generated gene expression levels according to ***m***_*j*_ = ***X***_eQTL,*j*_***δ***_*j*_ + ***ϵ***_*m,j*_ where 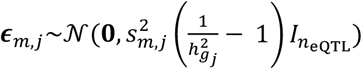 and 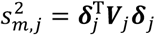. Last, we computed the marginal *cis*-eQTL effect sizes 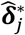 and corresponding statistics using simple linear regression, and we only kept the most significant association per gene.

We repeated this simulation pipeline for all *t* perturbed genes and concatenated all simulated results to get genome-wide genotype matrix ***X***_eQTL_ ∈ ℝ^*n*×*k*^, *cis*-eQTL true effect size matrix **Δ** ∈ ℝ^*k*×*t*^, *cis*-eQTL estimated effect size matrix 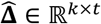 and LD matrix ***V*** ∈ ℝ^*k*×*k*^. For simplicity, we assume all genes have the same number of *cis*-eQTLs (*k*_*j*_; e.g., 1, 2 or 3), so the total number of *cis*-eQTLs is *k* = *tk*_*j*_. We guaranteed that the correlations between autosome SNPs are zero and the LD patterns within autosomes capture the correlation of real *cis*-eQTLs across genes.

### Simulating GWAS summary data

To reflect the realistic case that the GWAS and *cis*-eQTL summary data are obtained from different groups of individuals, we re-simulated centered and standardized genotype data 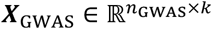 from 𝒩(**0, *V***) where *n*_GWAS_ ∈ {1 × 10^4^, 5 × 10^4^, 1 × 10^5^, 2 × 10^5^}. In our simulations, we directly utilized the real Perturb-seq effect size matrix from Yao et al.^25^ This matrix contains 600 (459 overlapped with eQTLGen) protein-coding perturbed genes and 13,151 downstream genes. To construct the perturbation vector 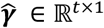, we first randomly selected one downstream gene to serve as the mediating gene. We then sampled *t* ∈ {200, 400, 600, 800} perturbed genes with replacement. Since sampling with replacement can result in duplicate effect sizes, we added small random noise drawn from a standard normal distribution to all values to ensure variability among the selected entries.

We simulated a complex trait vector 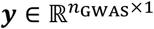, according to ***y*** = ***X***_GWAS_**Δ*γ****α* + ***ϵ*** where **Δ** is the *cis*-eQTL real effect size matrix, ***γ*** is the perturbation effect size vector, 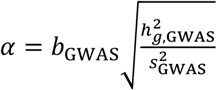 is the mediating effect size, *b*_GWAS_∼sample{−1, 1} is the mediating effect size sign, 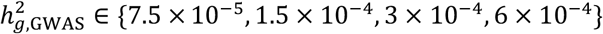 is the per-mediating-gene GWAS heritability (i.e., the proportion of variation of the complex trait explained by single mediating gene expression), 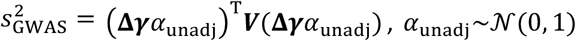 is the mediating effect unadjusted for heritability, and 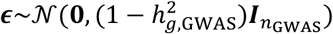. Last, we performed simple linear regression on ***y*** for each SNP in to compute the marginal effect size vector 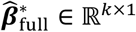 and its corresponding standard error diagonal matrix 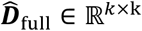. Because the Mr. PEG framework only focused on the top *cis*-eQTL for each perturbed gene, we reduced 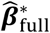 and 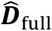 to 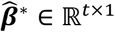 and 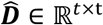, thus matching the entries in ***γ***. Overall, with marginal GWAS effect size 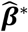, standard error 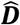, top marginal *cis*-eQTL effect size 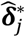, LD matrix ***V***, and perturb-seq vector ***γ***, we can infer Mr. PEG mediating effects 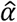.

We additionally simulated null GWAS data in the absence of any perturbation effect (**Fig. 1**). In this case, the *cis*-eQTLs affect the expression levels of their perturbed genes, and in turn influence the complex trait directly rather than via mediating genes. We computed the null complex trait ***y***_null_ = ***X***_GWAS_**Δ*ω*** + ***ϵ***_null_ where 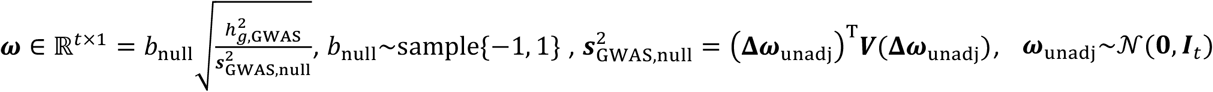, and 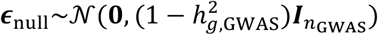. Then, we replaced ***y*** with ***y***_null_ and computed marginal GWAS effect size 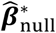 using linear regression. Last, we applied Mr. PEG using 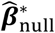, marginal *cis*-eQTL effect estimate 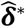, and perturbational effects 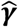 to test the non-existent mediating effects.

### Simulating model misspecifications

To evaluate the performance when there exists pleiotropy from SNPs to the complex traits other than perturbational mediating effects, we simulated the complex trait according to 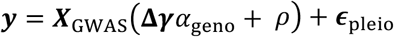 where 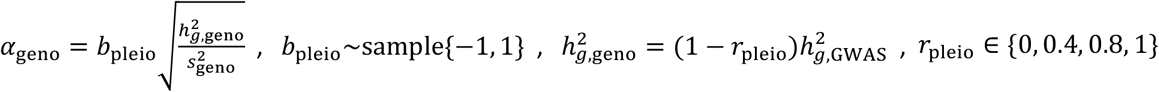 is the proportion of 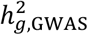 explained by pleiotropy, 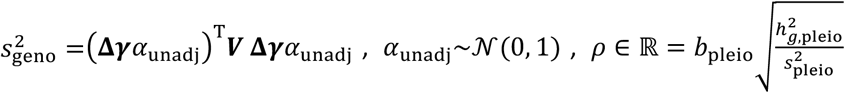 is the SNP pleiotropy effects, 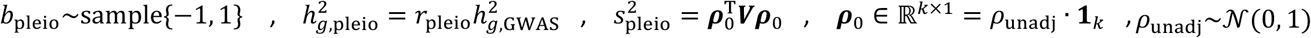, and 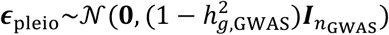. To evaluate the performance when the mediating genes’ own *cis*-eQTLs also have pleiotropy effects on the complex traits (indepedent from mediating effects *α*), we simulated the complex trait according to 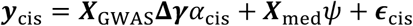 where 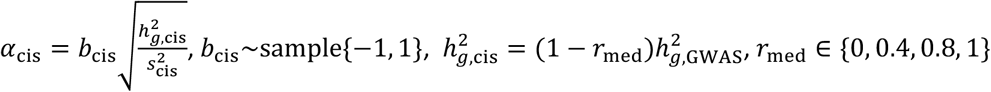 is the proportion of 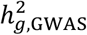 explained by mediating genes’ *cis*-eQTL, 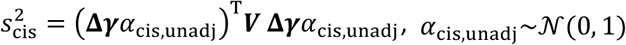, and 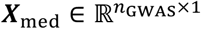 is the genotype for mediating genes. For simplicity, we assume each mediating gene only harbors one *cis*-eQTL. We simulated ***X***_med_ from 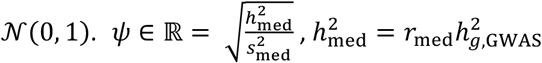, and 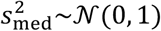. Perturbational experiments may include genes that are irrelevant to the complex traits, which can be seen as regulatory networks noise. Therefore, we additionally included *t*_add_ ∈ {200, 400, 800} more perturbed genes (and simulated other necessary parameters) in the Mr. PEG framework while they did not contribute to the complex traits. Conversely, the perturbed genes that function in the regulatory networks of the complex trait may not be available in practice. Therefore, we randomly selected *t*_test_ ∈ {50, 100, 200} out of the default number of perturbed genes (*t* = 400) and observed the performance difference. We also evaluated the cases when marginal *cis*-eQTL effect estimate 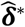 is missing in the inference but *cis*-eQTLs still influence perturbed genes, and in turn mediating genes and complex traits. We replaced 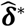 with 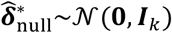, and applied Mr. PEG using 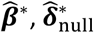, and perturbational effects 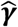. To evaluate the performance difference between using *cis*-eQTL effect estimates and statistics, we also replace *cis*-eQTL effect sizes with their corresponding t statistics in the inference.

### Simulation parameters

We repeated the simulation pipeline 500 times to evaluate the false positive rate and power. We compared the performance between four versions of Mr. PEG: (1) using standard MLE with its analytically derived standard error (denoted as “Standard”), (2) using MLE with standard error estimated from permutation tests (“Permutation”), and (3–4) using either an in-sample or an out-of-sample LD matrix to reflect practical scenarios. For the permutation test, we permutated the perturbational effects across genes for 500 times in the inference. That is, we kept SNP’s original GWAS effects, standard errors, and their *cis*-eQTL statistics but permutated the perturbational effects. We computed the LD matrix using genotypes from 398 individuals with European ancestry in TOPMed MESA study as out-of-sample LD. We varied five parameters: GWAS sample size (*n*_GWAS_), *cis*-eQTL sample size (*n*_eQTL_), the number of *cis*-eQTLs per mediating gene (*k*_*j*_), the number of perturbed genes (*t*), and GWAS heritability explained by one mediating gene 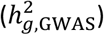. Unless stated otherwise, the default parameters are *n*_GWAS_ = 2 × 10^5^, *n*_eQTL_ = 200, *k*_*j*_ = 2, *t* = 400, and 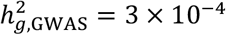. We used PLINK2^63,64^ to prepare genotypes for simulations. We also evaluated the unbiasedness of our estimator 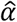 by computing its bias across simulations as 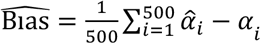 across simulation repetition.

### Perturbational screens, *cis*-eQTL, and GWAS data

For perturbational screening data, we obtained effect size estimates from Yao et al.^25^, which developed two experimental strategies to generate composite samples for Perturb-seq screens, substantially reducing experimental costs. They also introduced an inference method, FR-Perturb, designed to estimate gene-to-gene perturbation effect sizes. The key idea of FR-Perturb is to apply factor analysis to project the single-cell count matrix into a lower-dimensional space, and then infer the genetic perturbation effects on these lower-dimensional factors, thereby enhancing statistical power. The Perturb-seq effect size matrix we analyzed was computed on data aggregated from both the conventional and the compressed screening experiments using FR-Perturb with 10 latent factors. In total, the dataset from Yao et al. includes 600 upstream (perturbed) genes and 16,952 downstream genes. For GWAS data, we obtained summary statistics from 43 genome-wide association studies (GWAS), including 11 autoimmune diseases, 18 blood-related traits, and various other complex traits, with an average effective sample size of 228,500 (**Supplementary Table 1**). For *cis*-eQTL data, we used summary statistics from eQTLGen Phase I, based on whole blood samples from 31,684 individuals of European ancestry. This dataset includes 3,663,456 unique *cis*-eQTLs for 16,987 genes across the 22 autosomes.

We integrated these datasets as follows. In total, of the 600 perturbed genes in the Yao dataset, 458 are also present in the eQTLGen dataset. For downstream genes, we restricted our analysis to protein-coding genes located on autosomes, resulting in 13,151 downstream genes used in the Mr. PEG inference. We used two versions of the Perturb-seq effect size matrix in our analysis: (1) the full matrix, which includes all original effect size values; and (2) a sparse matrix, in which only the top 1% of absolute effect sizes are retained, and all remaining values are set to zero. To ensure the robustness of inference when using the sparse matrix, we limited our analysis to genes that had at least 10 upstream genes with non-zero entries. We ran Mr. PEG using LD-pruned *cis*-eQTLs: to better incorporate genes’ perturbation effects, we preferentially retained SNPs whose corresponding perturbed genes exert larger absolute effects on downstream genes. Specifically, we first ranked perturbed genes in descending order based on their average absolute effect sizes across all downstream genes. We then retained the top *cis*-eQTL of the highest-ranked perturbed gene and removed all other *cis*-eQTLs within a 1 Mb window that showed an absolute LD correlation greater than 0.1, along with their associated perturbed genes. This procedure was repeated iteratively for the next available gene in the ranked list until no perturbed genes remained. To compute the LD, we used genotypes from 489 individuals of European ancestry from the 1000 Genomes Project Phase 3.

To evaluate that our real-data analysis was controlled for false positives, we shuffled effect sizes across SNPs from inflammatory bowel disease (IBD) GWAS by de Lange et al.^38^, one of our 43 GWASs, and applied Mr. PEG with real *cis*-eQTL and Perturb-seq data. We applied both full and sparse Perturb-seq effect size matrices with LD-pruned and LD-adjusted inference, respectively, resulting in 8 sets of results. We determined Mr. PEG significant genes with adjusted P value <0.05 using Bonferroni correction with n=23,000, which is roughly the number of protein-coded genes.

### Comparisons with other methods

In our real data analysis, we compared the results from Mr. PEG to several benchmark methods, including Summary-based Mendelian Randomization (SMR)^8^, *trans*-SMR, *trans*-MR, GWAS closest-gene mapping, and burden tests. SMR identifies genes whose predicted expression levels are associated with complex traits by leveraging GWAS summary statistics and top *cis*-eQTLs. We performed SMR using software from the Yang lab (**Code Availability**), with the same GWAS and *cis*-eQTL datasets used in Mr. PEG. For *trans*-SMR, we substituted each gene’s top *cis*-eQTL with its top *trans*-eQTL from the eQTLGen consortium. We manually computed the chi-square statistic as 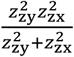 where 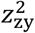 and 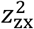 are the *Z* statistics from GWAS and *trans*-eQTL analysis, respectively, with 1 degree of freedom. For *trans*-MR, we applied traditional Mendelian randomization using all significant *trans*-eQTLs for each gene from eQTLGen, implemented via the TwoSampleMR R package^65^. We also curated the closest gene to each GWAS lead variant among the downstream genes tested by Mr. PEG. Specifically, we first identified all genome-wide significant SNPs and expanded their genomic regions by ±500 kb. We then merged overlapping windows and retained the most significant SNP within each clumped region. For each such region, we selected the gene closest to the lead SNP. Finally, we included genes identified through loss-of-function (LoF) burden tests in GENEBASS (**Data Availability**). For SMR, *trans*-SMR, and the burden test, we determined gene-level significance using a Bonferroni-corrected threshold of P<0.05/23,000. For *trans*-MR, due to its reduced statistical power, we reported genes with nominal P<0.05 without multiple testing correction.

### Gene annotations and other analysis

We obtained six metrics that quantify or correlate genes’ intolerance of loss-of-function mutation, including pLI^31^, LOEUF^32^, EDS^33^, s_het^34^, RVIS^35^, and hs^14,36^. For each metric, we performed a linear regression in which the dependent variable is the gene’s metric score and the independent variable is a binary indicator for whether the gene is Mr. PEG-significant, adjusted for three covariates: the indicator for full effect size matrix or sparse matrix, the number of non-zero perturbation effects, and the number of significant GWAS variants. To mitigate the influence of potential outliers, we performed permutation tests to recompute the standard errors of the linear regression estimates and the corresponding P values.

Specifically, we shuffled the binary indicator for whether a gene is Mr. PEG–significant across 100 iterations to construct an empirical distribution of the regression estimates, and used its standard deviation as the standard error.

We obtained gene-level annotations curated by Mostafavi et al.^14^ and compared several features, including “ABC_length_per_type,” “Roadmap_length_per_type,” “connect_decile,” and “hs.” Specifically, “ABC_length_per_type” is defined as the mean length of ABC enhancers across active biosamples, based on data processed from Nasser et al.;^40^ “Roadmap_length_per_type” represents the mean length of Roadmap Epigenomics Consortium enhancers across active biosamples, using data from Liu et al.;^66^ “connect_decile” refers to the quintile of the connectedness score computed from co-expression networks inferred by Saha et al.;^67^ “hs” corresponds to selection coefficient estimates from Agarwal et al.^36^ For each method (Mr. PEG, SMR, *trans*-SMR, *trans*-MR, Burden, and GWAS closest genes), we computed the average of these annotation values across their prioritized genes. To estimate standard errors while reducing the influence of outlier genes, we applied a bootstrap approach with 100 resampling iterations.

To ensure the relevance of the Perturb-seq experimental context (i.e., macrophage cell lines) to the complex traits analyzed, we selected the top 500 most highly expressed downstream genes in control cells (i.e., without perturbation) and mapped them to the genome to construct a gene-based annotation. We then ran stratified LD score regression (LDSC)^68^ to compute the enrichment.

We conducted gene set enrichment analysis (GSEA) using Gene Ontology Biological Process (GO BP) terms using clusterProfiler.^69^ All tested genes, along with their corresponding test statistics, were used as input annotations. We considered BP gene sets ranging in size from 10 to 17,000 genes. For Mr. PEG, we assigned each gene the maximum Z statistics obtained from the full and sparse versions of the Perturb-seq effect size matrix. For GWAS-based annotations, each gene was assigned the Z statistics of the most significant SNP located within a 1 Mb window surrounding the gene. To ensure consistency in interpretation across methods, we converted all test statistics to their absolute values, such that a positive enrichment score indicates enrichment, and a negative score indicates depletion. We also performed GSEA using Mr. PEG with the original (non–absolute) test statistics to examine differences between upregulated and downregulated genes associated with increased disease risk.

To assess the relationship between Mr. PEG and other methods’ effect sizes with loss-of-function (LoF) burden effect sizes, we conducted linear regression analyses concatenating results across all the complex traits. The dependent variable was the LoF burden effect size from GENEBASE (**Data Availability**), and the independent variable was the effect size estimated by each method. Covariates include the dummy variables for complex traits.

To assess the enrichment of Mr. PEG genes using external datasets, we first obtained the Inborn Errors of Immunity (IEI) gene list^44^ and the gene sets derived from STING-seq^45^. For STING-seq, we selected cis-targeted genes across all blood traits that were significant (q < 0.05) within a ±500 kb window, based on GWAS conducted in individuals of European ancestry. For each method, among all tested candidate genes, we performed logistic regression with a binary dependent variable indicating whether a gene belongs to the IEI gene set, and a binary independent variable indicating whether the gene was identified as significant by that method. Finally, we meta-analyzed the regression results across all traits for the IEI gene set, and across all blood-related traits for the STING-seq gene set.

In our case study, we assessed the GWAS significance of lead SNPs associated with Mr. PEG-prioritized genes by comparing the GWAS data we used with larger, more recent meta-analyses from the VA Million Veteran Program (MVP), FinnGen, and UK Biobank (UKBB) (**Data Availability**). To explore potential causal relationships between complex traits and drug usage, we applied Generalized Summary-based Mendelian Randomization^52,53^ (GSMR) across our 43 GWASs. Drug usage GWAS summary statistics were obtained from Wu et al.^54^

## Data Availability

All data produced are available online at https://zenodo.org/records/17907323

## Acknowledgements

The authors thank Dr. Brian Cleary of Boston University for his valuable help on this manuscript. The authors thank members of the Gusev Lab at Dana-Farber Cancer Institute and Harvard Medical School, the Mancuso Lab at University of Southern California (USC), and the Gazal Lab at USC for fruitful discussions regarding this manuscript.

## Data availability

All our analysis data is available at https://zenodo.org/records/17907323

genomAD: https://gnomad.broadinstitute.org

FINNGEN+MVP+UKBB GWAS: https://mvp-ukbb.finngen.fi

GENEBASS: http://genebass.org

KEGG pathway lookup: https://www.genome.jp/kegg/pathway.html

## Code availability

Our analysis codes are available at https://github.com/gusevlab/mrpeg_analysis_codes_public

Mr. PEG software is available at https://github.com/gusevlab/mrpeg

SMR: https://yanglab.westlake.edu.cn/software/smr/#Overview

TWOSampleMR: https://github.com/MRCIEU/TwoSampleMR

clusterProfiler: https://guangchuangyu.github.io/software/clusterProfiler/

LDSC: https://github.com/bulik/ldsc

