## Supplemental Note for "Integrating perturbational screens, eQTL, and GWAS data identifies mediating genes for complex traits"

### Supplementary Note

#### 1 Generative Model

Mr. PEG is short for Mendelian Randomization integrating perturbational screens, eQTL, and GWAS summary data. In this section, we provide a detailed description for the generative model of Mr. PEG framework.

In this study, a perturbation refers to a CRISPR-based experimental intervention that knocks out one or more genes (referred to as perturbed genes or upstream genes). Following a perturbation, the expression levels of other genes (referred to as mediating genes or downstream genes) are measured using scRNA-seq or similar techniques.

Mr. PEG models a complex trait as a function of the expression levels of some mediating gene, which is further modeled as a linear combination of the expression levels of perturbed genes. Finally, the expression levels of perturbed genes are modeled as a linear combination of genotyped variants. This hierarchical framework captures the cascading relationships between eQTLs, perturbed genes, and mediating genes, ultimately linking genetic variation to complex traits. We will define this in mathematical equations step by step. We also assume complex traits, gene expression levels, and genotypes are centered at zero and scaled by their standard deviations.

First, we model the complex trait $\mathbf{y} \in \mathbb{R}^{n\times 1}$ for $n$ individuals as a function of a focal gene expression level vector $\mathbf{g} \in \mathbb{R}^{n\times 1}$ with effect size scalar $\alpha \in \mathbb{R}$ and the environmental noise vector $\boldsymbol{\epsilon}_y \in \mathbb{R}^{n\times 1} \sim \mathcal{N}(\mathbf{0}, \sigma_\epsilon^2 \mathbf{I}_{n\times n})$. The equation follows as:

$$\mathbf{y} = \mathbf{g}\alpha + \boldsymbol{\epsilon}_y. \tag{1}$$

Next, we further model $\mathbf{g}$ as a linear combination of $t$ perturbed gene expression level matrix $\mathbf{M} \in \mathbb{R}^{n\times t}$ with a gene-to-gene effect size vector $\boldsymbol{\gamma} \in \mathbb{R}^{t\times 1}$ and the environmental noise vector $\boldsymbol{\epsilon}_g \in \mathbb{R}^{n\times 1} \sim \mathcal{N}(\mathbf{0}, \sigma_g^2 \mathbf{I}_{n\times n})$ as

$$\mathbf{g} = \mathbf{M}\boldsymbol{\gamma} + \boldsymbol{\epsilon}_g = \sum_{j=1}^{t} \mathbf{m}_j \gamma_j + \boldsymbol{\epsilon}_g. \tag{2}$$

where $j$ is the perturbed gene index, $\mathbf{m}_j \in \mathbb{R}^{n\times 1}$ is the corresponding gene expression level vector of a perturbed gen, and $\gamma_j \in \mathbb{R}$ is the effect size scalar of a perturbed gene on the focal gene.

Last, we model $\mathbf{m}_j$ as a linear combination of the genotype matrix $\mathbf{X}_j \in \mathbb{R}^{n\times k_j}$ with the eQTL effect size vector $\boldsymbol{\delta}_j \in \mathbb{R}^{k_j\times 1}$ and the environmental noise vector $\boldsymbol{\epsilon}_m \in \mathbb{R}^{n\times 1} \sim \mathcal{N}(\mathbf{0}, \sigma_m^2 \mathbf{I}_{n\times n})$ where $k_j$ is the number of SNPs for the perturbed gene. The equation follows as:

$$\mathbf{m}_j = \mathbf{X}_j \boldsymbol{\delta}_j + \boldsymbol{\epsilon}_m. \tag{3}$$

The algebraic expansion after plugging 2 and 3 into 1, we have:

$$\begin{aligned}
\mathbf{y} &= \mathbf{g}\alpha + \boldsymbol{\epsilon}_y \\
&= (\mathbf{M}\boldsymbol{\gamma} + \boldsymbol{\epsilon}_g)\alpha + \boldsymbol{\epsilon}_y \\
&= (\sum_{j=1}^{t} \mathbf{m}_j \gamma_j + \boldsymbol{\epsilon}_g)\alpha + \boldsymbol{\epsilon}_y \\
&= \left[\sum_{j=1}^{t} (\mathbf{X}_j \boldsymbol{\delta}_j + \boldsymbol{\epsilon}_m)\gamma_j + \boldsymbol{\epsilon}_g\right]\alpha + \boldsymbol{\epsilon}_y \\
&= \sum_{j=1}^{t} \mathbf{X}_j \boldsymbol{\delta}_j \gamma_j \alpha + \boldsymbol{\epsilon}
\end{aligned}$$

where the last term holds as we push all the environmental noises into one environmental noise vector $\boldsymbol{\epsilon} \in \mathbb{R}^{n\times 1}$.

We can re-write $\sum_{j=1}^{t} (\mathbf{X}_j \boldsymbol{\delta}_j)\gamma_j = \mathbf{X}\boldsymbol{\Delta}\boldsymbol{\gamma}$ where $\mathbf{X} \in \mathbb{R}^{n\times k}$, $k$ is the number of unique SNPs, and $\boldsymbol{\Delta} \in \mathbb{R}^{k\times t}$ is the eQTL effect matrix of $k$ SNPs to $t$ perturbed genes. Note that if there are no overlapped SNPs across $t$ perturbed genes, then $k = \sum_{j=1}^{t} k_j$. In general, $\boldsymbol{\Delta}$ is sparse because each gene (i.e., a column in $\boldsymbol{\Delta}$) only has a few eQTLs (i.e., non-zero entries), thus the rest entries in the column are zero. $k = t$ suggests that each perturbed gene only harbors one eQTL, so $\boldsymbol{\Delta}$ is reduced to a diagonal matrix. Plugging in the new notation leads to:

$$\begin{aligned}
\mathbf{y} &= \mathbf{X}\underbrace{\boldsymbol{\Delta}\boldsymbol{\gamma}\alpha}_{\boldsymbol{\beta}} + \boldsymbol{\epsilon} \\
&= \mathbf{X}\boldsymbol{\beta} + \boldsymbol{\epsilon}
\end{aligned} \tag{4}$$

where $\boldsymbol{\beta} \in \mathbb{R}^{p\times 1}$ is the SNP effect size on the trait.

We also note that $\boldsymbol{\Delta}$ can be any *cis* or *trans* molecular QTL if the molecular feature of a gene influences the expression of downstream genes. We use *cis*-eQTLs, as they are the most commonly studied type of molecular QTL. Similarly, $\gamma$ can be any gene-to-gene regulatory effect (not necessarily those derived from perturbational experiments) if it represents how an upstream gene's expression affects the expression of downstream genes. Here, we use effects estimated from CRISPR-based perturbational screens, because these experiments provide causal evidence of regulatory relationships, rather than correlation.

#### 2 Inference

Let $\hat{\boldsymbol{\beta}}^*$ be the marginal effect size estimate for eash SNP in GWAS and we assume $\hat{\boldsymbol{\beta}}^* \sim \mathcal{N}(\hat{\mathbf{V}}\boldsymbol{\beta}, \sigma^2\hat{\mathbf{D}}\hat{\mathbf{V}}\hat{\mathbf{D}})$ where $\hat{\mathbf{D}} \in \mathbb{R}^{k\times k}$ is a diagonal matrix where the entries are the standard error of $\hat{\boldsymbol{\beta}}^*$ for each SNP, $\hat{\mathbf{V}} \in \mathbb{R}^{k\times k}$ is the estimated SNP correlation matrix (i.e., LD matrix; usual), and $\sigma^2 \in \mathbb{R}_+$ is the heterogeneity parameter.

Based on 4, we have:

$$\hat{\boldsymbol{\beta}}^* \sim \mathcal{N}(\alpha\hat{\mathbf{V}}\boldsymbol{\Delta}\gamma, \sigma^2\hat{\mathbf{D}}\hat{\mathbf{V}}\hat{\mathbf{D}}). \tag{5}$$

Therefore, the log-likelihood for $\alpha$ and $\sigma^2$ is:

$$\begin{aligned}
\ell(\alpha, \sigma^2 \mid \hat{\boldsymbol{\beta}}^*, \boldsymbol{\Delta}, \gamma, \hat{\mathbf{D}}, \hat{\mathbf{V}}) &= \log \mathcal{N}(\hat{\boldsymbol{\beta}}^* \mid \boldsymbol{\Delta}, \gamma, \alpha, \sigma^2, \hat{\mathbf{D}}, \hat{\mathbf{V}}) \\
&= \log \frac{1}{(2\pi)^{k/2}(|\sigma^2\hat{\mathbf{D}}\hat{\mathbf{V}}\hat{\mathbf{D}}|)^{k/2}} \exp\left(-\frac{1}{2}(\boldsymbol{\beta}^* - \alpha\hat{\mathbf{V}}\boldsymbol{\Delta}\gamma)^\intercal(\sigma^2\hat{\mathbf{D}}\hat{\mathbf{V}}\hat{\mathbf{D}})^{-1}(\hat{\boldsymbol{\beta}}^* - \alpha\hat{\mathbf{V}}\boldsymbol{\Delta}\gamma)\right) \\
&= \log \frac{1}{(2\pi)^{k/2}(\sigma^2|\hat{\mathbf{D}}\hat{\mathbf{V}}\hat{\mathbf{D}}|)^{k/2}} \exp\left(-\frac{1}{2}(\hat{\boldsymbol{\beta}}^* - \alpha\hat{\mathbf{V}}\boldsymbol{\Delta}\gamma)^\intercal(\sigma^2\hat{\mathbf{D}}\hat{\mathbf{V}}\hat{\mathbf{D}})^{-1}(\hat{\boldsymbol{\beta}}^* - \alpha\hat{\mathbf{V}}\boldsymbol{\Delta}\gamma)\right) \\
&= -\frac{k}{2}\log(2\pi) - \frac{k}{2}\log\left(\sigma^2|\hat{\mathbf{D}}\hat{\mathbf{V}}\hat{\mathbf{D}}|\right) - \frac{1}{2}(\hat{\boldsymbol{\beta}}^* - \alpha\hat{\mathbf{V}}\boldsymbol{\Delta}\gamma)^\intercal(\sigma^2\hat{\mathbf{D}}\hat{\mathbf{V}}\hat{\mathbf{D}})^{-1}(\hat{\boldsymbol{\beta}}^* - \alpha\hat{\mathbf{V}}\boldsymbol{\Delta}\gamma)
\end{aligned}$$

Then, the maximumn likelihood estimators (MLE) for $\alpha$ and $\sigma^2$ are:

$$\begin{aligned}
\hat{\alpha} &= \arg\max_{\alpha} \ell(\alpha \mid \sigma^2, \hat{\boldsymbol{\beta}}^*, \boldsymbol{\Delta}, \gamma, \hat{\mathbf{D}}, \hat{\mathbf{V}}) \\
\hat{\sigma}^2 &= \arg\max_{\sigma^2} \ell(\sigma^2 \mid \alpha, \boldsymbol{\beta}^*, \boldsymbol{\Delta}, \gamma, \hat{\mathbf{D}}, \hat{\mathbf{V}})
\end{aligned}$$

As a result, the MLE of $\alpha$ is the solution to $\frac{\partial\ell}{\partial\alpha} = 0$:

$$
\begin{aligned}
\frac{\partial \ell}{\partial \alpha} &= \frac{1}{2}\left[(\hat{\boldsymbol{\beta}}^* - \alpha\hat{\mathbf{V}}\boldsymbol{\Delta}\gamma)^\intercal \left(\sigma^2\hat{\mathbf{D}}\hat{\mathbf{V}}\hat{\mathbf{D}}\right)^{-1}\hat{\mathbf{V}}\boldsymbol{\Delta}\gamma + (\hat{\boldsymbol{\beta}}^* - \alpha\hat{\mathbf{V}}\boldsymbol{\Delta}\gamma)^\intercal \left(\sigma^2\hat{\mathbf{D}}\hat{\mathbf{V}}\hat{\mathbf{D}}\right)^{-1}\hat{\mathbf{V}}\boldsymbol{\Delta}\gamma\right] \\
&= (\hat{\boldsymbol{\beta}}^* - \alpha\hat{\mathbf{V}}\boldsymbol{\Delta}\gamma)^\intercal(\sigma^2\hat{\mathbf{D}}\hat{\mathbf{V}}\hat{\mathbf{D}})^{-1}\hat{\mathbf{V}}\boldsymbol{\Delta}\gamma \\
&= 0
\end{aligned}
$$

$$
\begin{aligned}
\hat{\alpha} &= \left[(\hat{\mathbf{V}}\boldsymbol{\Delta}\gamma)^\intercal(\hat{\mathbf{D}}\hat{\mathbf{V}}\hat{\mathbf{D}})^{-1}\hat{\mathbf{V}}\boldsymbol{\Delta}\gamma\right]^{-1}(\hat{\mathbf{V}}\boldsymbol{\Delta}\gamma)^\intercal(\hat{\mathbf{D}}\hat{\mathbf{V}}\hat{\mathbf{D}})^{-1}\hat{\boldsymbol{\beta}}^* \\
&= \frac{(\hat{\mathbf{V}}\boldsymbol{\Delta}\gamma)^\intercal(\hat{\mathbf{D}}\hat{\mathbf{V}}\hat{\mathbf{D}})^{-1}\hat{\boldsymbol{\beta}}^*}{(\hat{\mathbf{V}}\boldsymbol{\Delta}\gamma)^\intercal(\hat{\mathbf{D}}\hat{\mathbf{V}}\hat{\mathbf{D}})^{-1}\hat{\mathbf{V}}\boldsymbol{\Delta}\gamma}
\end{aligned}
$$

We assume $\boldsymbol{\Delta} = \hat{\mathbf{V}}^{-1}\hat{\boldsymbol{\Delta}}^*$ in a well-powered *cis*-eQTL study (e.g., eQTLGen) where $\hat{\boldsymbol{\Delta}}^*$ is the marginal *cis*-eQTL effect size estimates. $\hat{\mathbf{V}}$ is typically obtained from external reference genotype data. We heuristically assume $\boldsymbol{\gamma} = \hat{\gamma}$ in well-designed experiments. We note that the uncertainty of the estimates $\hat{\boldsymbol{\Delta}}^*$ and $\hat{\gamma}$ is not modeled. For $\hat{\boldsymbol{\Delta}}^*$, we can instead use the corresponding Z statistics to incorporate uncertainty, and this is less of an issue in a well-powered *cis*-eQTL study. For $\hat{\gamma}$, many methods rely on permutation tests to obtain P values because operturbational effect estimates do not follow a normal distribution. Therefoere, Z statistics derived from discrete P values in permutation test may not perform well in this setting.

Therefore, we express the maximum likelihood estimate (MLE) for $\alpha$ using these practical estimates as:

$$
\hat{\alpha} = \frac{(\hat{\boldsymbol{\Delta}}^*\hat{\gamma})^\intercal(\hat{\mathbf{D}}\hat{\mathbf{V}}\hat{\mathbf{D}})^{-1}\hat{\boldsymbol{\beta}}^*}{(\hat{\boldsymbol{\Delta}}^*\hat{\gamma})^\intercal(\hat{\mathbf{D}}\hat{\mathbf{V}}\hat{\mathbf{D}})^{-1}(\hat{\boldsymbol{\Delta}}^*\hat{\gamma})} \tag{6}
$$

Similarly, the MLE of $\sigma^2$ is the solution to $\frac{\partial \ell}{\partial \sigma^2} = 0$:

$$
\begin{aligned}
\frac{\partial \ell}{\partial \sigma^2} &= -\frac{k}{2\sigma^2} + \frac{1}{2}(\hat{\boldsymbol{\beta}}^* - \alpha\hat{\mathbf{V}}\boldsymbol{\Delta}\gamma)^\intercal(\sigma^2\hat{\mathbf{D}}\hat{\mathbf{V}}\hat{\mathbf{D}})^{-1}\hat{\mathbf{D}}\hat{\mathbf{V}}\hat{\mathbf{D}}(\sigma^2\hat{\mathbf{D}}\hat{\mathbf{V}}\hat{\mathbf{D}})^{-1}(\hat{\boldsymbol{\beta}}^* - \alpha\hat{\mathbf{V}}\boldsymbol{\Delta}\gamma) \\
&= -\frac{k}{2\sigma^2} + \frac{1}{2\sigma^4}(\hat{\boldsymbol{\beta}}^* - \alpha\hat{\mathbf{V}}\boldsymbol{\Delta}\gamma)^\intercal(\hat{\mathbf{D}}\hat{\mathbf{V}}\hat{\mathbf{D}})^{-1}\hat{\mathbf{D}}\hat{\mathbf{V}}\hat{\mathbf{D}}(\hat{\mathbf{D}}\hat{\mathbf{V}}\hat{\mathbf{D}})^{-1}(\hat{\boldsymbol{\beta}}^* - \alpha\hat{\mathbf{V}}\boldsymbol{\Delta}\gamma) \\
&= -\frac{k}{2\sigma^2} + \frac{1}{2\sigma^4}(\hat{\boldsymbol{\beta}}^* - \alpha\hat{\mathbf{V}}\boldsymbol{\Delta}\gamma)^\intercal(\hat{\mathbf{D}}\hat{\mathbf{V}}\hat{\mathbf{D}})^{-1}(\hat{\boldsymbol{\beta}}^* - \alpha\hat{\mathbf{V}}\boldsymbol{\Delta}\gamma) \\
&= 0 \\
\hat{\sigma}^2_{\text{MLE}} &= \frac{1}{k}(\hat{\boldsymbol{\beta}}^* - \hat{\alpha}\hat{\boldsymbol{\Delta}}^*\hat{\gamma})^\intercal(\hat{\mathbf{D}}\hat{\mathbf{V}}\hat{\mathbf{D}})^{-1}(\hat{\boldsymbol{\beta}}^* - \hat{\alpha}\hat{\boldsymbol{\Delta}}^*\hat{\gamma})
\end{aligned}
$$

We also note that the unbiased estimator for $\hat{\sigma}$ is

$$\hat{\sigma}^2_{\text{UB}} = \frac{1}{k-1}(\hat{\boldsymbol{\beta}}^* - \hat{\alpha}\hat{\boldsymbol{\Delta}}^*\hat{\gamma})^\intercal(\hat{\mathbf{D}}\hat{\mathbf{V}}\hat{\mathbf{D}})^{-1}(\hat{\boldsymbol{\beta}}^* - \hat{\alpha}\hat{\boldsymbol{\Delta}}^*\hat{\gamma}) \tag{7}$$

Next, the variance of $\hat{\alpha}$ is

$$\begin{aligned}
\mathbb{V}(\hat{\alpha}) &= \mathbb{V}\left[\frac{(\hat{\mathbf{V}}\boldsymbol{\Delta}\gamma)^\intercal(\hat{\mathbf{D}}\hat{\mathbf{V}}\hat{\mathbf{D}})^{-1}\hat{\boldsymbol{\beta}}^*}{(\hat{\mathbf{V}}\boldsymbol{\Delta}\gamma)^\intercal(\hat{\mathbf{D}}\hat{\mathbf{V}}\hat{\mathbf{D}})^{-1}(\hat{\mathbf{V}}\boldsymbol{\Delta}\gamma)}\right] \\
&= \frac{(\hat{\mathbf{V}}\boldsymbol{\Delta}\gamma)^\intercal(\hat{\mathbf{D}}\hat{\mathbf{V}}\hat{\mathbf{D}})^{-1}\mathbb{V}\left[\hat{\boldsymbol{\beta}}^*\right]\left((\hat{\mathbf{V}}\boldsymbol{\Delta}\gamma)^\intercal(\hat{\mathbf{D}}\hat{\mathbf{V}}\hat{\mathbf{D}})^{-1}\right)^\intercal}{\left[(\hat{\mathbf{V}}\boldsymbol{\Delta}\gamma)^\intercal(\hat{\mathbf{D}}\hat{\mathbf{V}}\hat{\mathbf{D}})^{-1}(\hat{\mathbf{V}}\boldsymbol{\Delta}\gamma)\right]^2} \\
&= \frac{\sigma^2(\hat{\mathbf{V}}\boldsymbol{\Delta}\gamma)^\intercal(\hat{\mathbf{D}}\hat{\mathbf{V}}\hat{\mathbf{D}})^{-1}(\hat{\mathbf{D}}\hat{\mathbf{V}}\hat{\mathbf{D}})(\hat{\mathbf{D}}\hat{\mathbf{V}}\hat{\mathbf{D}})^{-1}(\hat{\mathbf{V}}\boldsymbol{\Delta}\gamma)}{\left[(\hat{\mathbf{V}}\boldsymbol{\Delta}\gamma)^\intercal(\hat{\mathbf{D}}\hat{\mathbf{V}}\hat{\mathbf{D}})^{-1}(\hat{\mathbf{V}}\boldsymbol{\Delta}\gamma)\right]^2} \\
&= \frac{\sigma^2}{(\hat{\mathbf{V}}\boldsymbol{\Delta}\gamma)^\intercal(\hat{\mathbf{D}}\hat{\mathbf{V}}\hat{\mathbf{D}})^{-1}(\hat{\mathbf{V}}\boldsymbol{\Delta}\gamma)}
\end{aligned}$$

The test statistic for $\hat{\alpha}$ will be :

$$\begin{aligned}
z_{\hat{\alpha}} &= \frac{\hat{\alpha} - 0}{\sqrt{\mathbb{V}(\hat{\alpha})}} \\
&= \frac{(\hat{\boldsymbol{\Delta}}^*\hat{\gamma})^\intercal(\hat{\mathbf{D}}\hat{\mathbf{V}}\hat{\mathbf{D}})^{-1}\hat{\boldsymbol{\beta}}^*}{\hat{\sigma}^2_{\text{UB}}\sqrt{(\hat{\boldsymbol{\Delta}}^*\hat{\gamma})^\intercal(\hat{\mathbf{D}}\hat{\mathbf{V}}\hat{\mathbf{D}})^{-1}(\hat{\boldsymbol{\Delta}}^*\hat{\gamma})}}.
\end{aligned}$$

The test statistic follows a $t$ distribution with $k-1$ degrees of freedom.

Alternatively, we can perform a permutation test to construct the null distribution for $\hat{\alpha}$ and compute its $\mu_{\hat{\alpha}}$ and $\text{SD}_{\hat{\alpha}}$. In detail, we shuffle only the rows in the perturbational effect estimates $\hat{\gamma}$ while keeping the SNP matching across *cis*-eQTL data, GWAS data, and the correlation matrix. The constructed null distribution assumes no association between complex traits and mediating genes, meaning any observed relationship is due solely to random perturbation effects rather than regulation by the perturbed genes and their *cis*-eQTLs. Last, when the number of perturbed genes in the model is large (e.g., $k > 100$), we can compute the permutation $Z$ statistic as $\frac{\hat{\alpha}-\mu_{\hat{\alpha}}}{\text{SD}_{\hat{\alpha}}}$, from which a two-sided P value based on the standard normal distribution can be obtained. Alternatively, when $k$ is small, the distribution of $\hat{\alpha}$ may deviate from normality; as a result, a discrete permutation-based $P$ value is preferred instead of computing a $Z$ statistic.

To make inference fast and stable, we can simplify our model as follows:

1. Because the most significant *cis*-eQTL (top *cis*-eQTL) of a gene usually explains the majority of ex-

pression signals, we can select a single top *cis*-eQTL for each perturbed gene. That is, $\hat{\mathbf{\Delta}}$ is reduced to a diagonal matrix with top *cis*-eQTL effect sizes with a dimension of $k \times k$ or $t \times t$ as $k = t$, so $\hat{\mathbf{\Delta}}\hat{\boldsymbol{\gamma}} = \hat{\boldsymbol{\delta}} \times \hat{\boldsymbol{\gamma}}$ where $\times$ means element-wise multiplication.

2. In practice, we also allow the corresponding marginal $Z$ statistics as replacements for marginal effect size $\hat{\mathbf{\Delta}}^*$ when it is not available.

3. If we perform LD pruning on SNPs before the inference such that top *cis*-eQTLs are independent, $\hat{\mathbf{V}}$ will become identity matrix, and further simply the model.

In addition, following previous work [1], we can assume a more complex model $\boldsymbol{\beta}^* \sim \mathcal{N}(\hat{\mathbf{D}}\hat{\mathbf{V}}\hat{\mathbf{D}}^{-1}\boldsymbol{\beta}, \sigma^2\hat{\mathbf{D}}\hat{\mathbf{V}}\hat{\mathbf{D}})$, the test statistics become:

$$z_{\hat{\alpha}} = \frac{(\hat{\mathbf{\Delta}}^*\hat{\gamma})^\intercal(\hat{\mathbf{D}}\hat{\mathbf{D}}\hat{\mathbf{V}})^{-1}\hat{\boldsymbol{\beta}}^*}{\hat{\sigma}^2_{\text{MLE}}\sqrt{(\hat{\mathbf{\Delta}}^*\hat{\gamma})^\intercal(\hat{\mathbf{V}}\hat{\mathbf{D}}\hat{\mathbf{V}}^{-1}\hat{\mathbf{D}}\hat{\mathbf{V}})^{-1}(\hat{\mathbf{\Delta}}^*\hat{\gamma})}}, \text{and}$$

$$\hat{\sigma}^2_{\text{MLE}} = \frac{1}{k}(\hat{\boldsymbol{\beta}}^* - \hat{\alpha}\hat{\mathbf{D}}\hat{\mathbf{V}}\hat{\mathbf{D}}^{-1}\hat{\mathbf{V}}^{-1}\hat{\mathbf{\Delta}}^*\hat{\gamma})^\intercal(\hat{\mathbf{D}}\hat{\mathbf{V}}\hat{\mathbf{D}})^{-1}(\hat{\boldsymbol{\beta}}^* - \hat{\alpha}\hat{\mathbf{D}}\hat{\mathbf{V}}\hat{\mathbf{D}}^{-1}\hat{\mathbf{V}}^{-1}\hat{\mathbf{\Delta}}^*\hat{\gamma})$$
