## Supplemental Figures for "Integrating perturbational screens, eQTL, and GWAS data identifies mediating genes for complex traits"

### Supplementary Figures

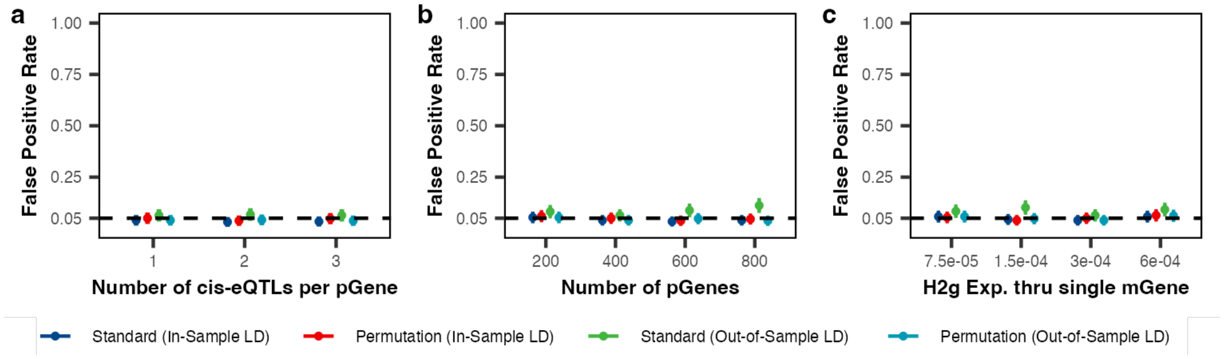


**Supplementary Figure 1: Mr. PEG controls the false positive rate at 5% in the absence of perturbation effects across different simulation parameters.**

(a) Number of *cis*-eQTLs per perturbed gene, with gene expression *cis*-SNP heritability held constant. (b) Number of perturbed genes affecting the mediating gene, where a larger number reflects more complex regulatory mechanisms. (c) Trait heritability (h2g) explained by a single mediating gene. Here, “pGene” refers to a perturbed gene and “mGene” refers to a mediating gene. We compared four versions of Mr. PEG: (1) using standard MLE with its analytically derived standard error (“Standard”), (2) using MLE with standard error estimated from permutation tests (“Permutation”), and (3–4) using either an in-sample or an out-of-sample LD matrix (**Methods**). We simulated scenarios under the null (no perturbation effects; “Null” in **Fig. 1**). Each parameter setting was evaluated using 500 independent replicates. The false positive rate was calculated as the proportion of replicates (out of 500) with a P value < 0.05 under the null scenario. Unless otherwise specified, the default parameters were: GWAS sample size = 100,000; *cis*-eQTL sample size = 200; number of *cis*-eQTLs = 1; number of perturbed genes contributing to the complex trait = 400; number of perturbed genes tested in inference = 400; per-gene trait heritability = $3 \times 10^{-4}$; no pleiotropic effects; and the mediating gene has no *cis*-eQTLs. The points are the mean across simulations, and the error bars are their corresponding 95% confidence intervals.

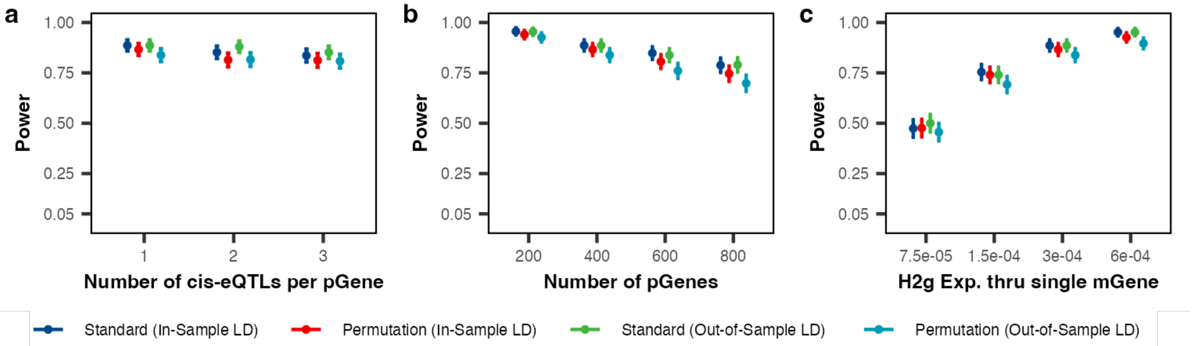


**Supplementary Figure 2: Mr. PEG achieves high power in the presence of perturbation effects across different simulation parameters.**

(a) Number of *cis*-eQTLs per perturbed gene, with gene expression *cis*-SNP heritability held constant. (b) Number of perturbed genes affecting the mediating gene, where a larger number reflects more complex regulatory mechanisms. (c) Trait heritability (h2g) explained by a single mediating gene. Here, "pGene" refers to a perturbed gene and "mGene" refers to a mediating gene. We compared four versions of Mr. PEG: (1) using standard MLE with its analytically derived standard error ("Standard"), (2) using MLE with standard error estimated from permutation tests ("Permutation"), and (3–4) using either an in-sample or an out-of-sample LD matrix (see **Methods**). We simulated scenarios under the alternative (with perturbation effects; "Alternative" in **Fig. 1**). Each parameter setting was evaluated using 500 independent replicates. Power was calculated as the proportion of replicates (out of 500) with a P value < 0.05 under the alternative scenario. Unless otherwise specified, the default parameters were: GWAS sample size = 100,000; *cis*-eQTL sample size = 200; number of *cis*-eQTLs = 1; number of perturbed genes contributing to the complex trait = 400; number of perturbed genes tested in inference = 400; per-gene trait heritability = $3 \times 10^{-4}$; no pleiotropic effects; and the mediating gene has no *cis*-eQTLs. The points are the mean across simulations, and the error bars are their corresponding 95% confidence intervals.

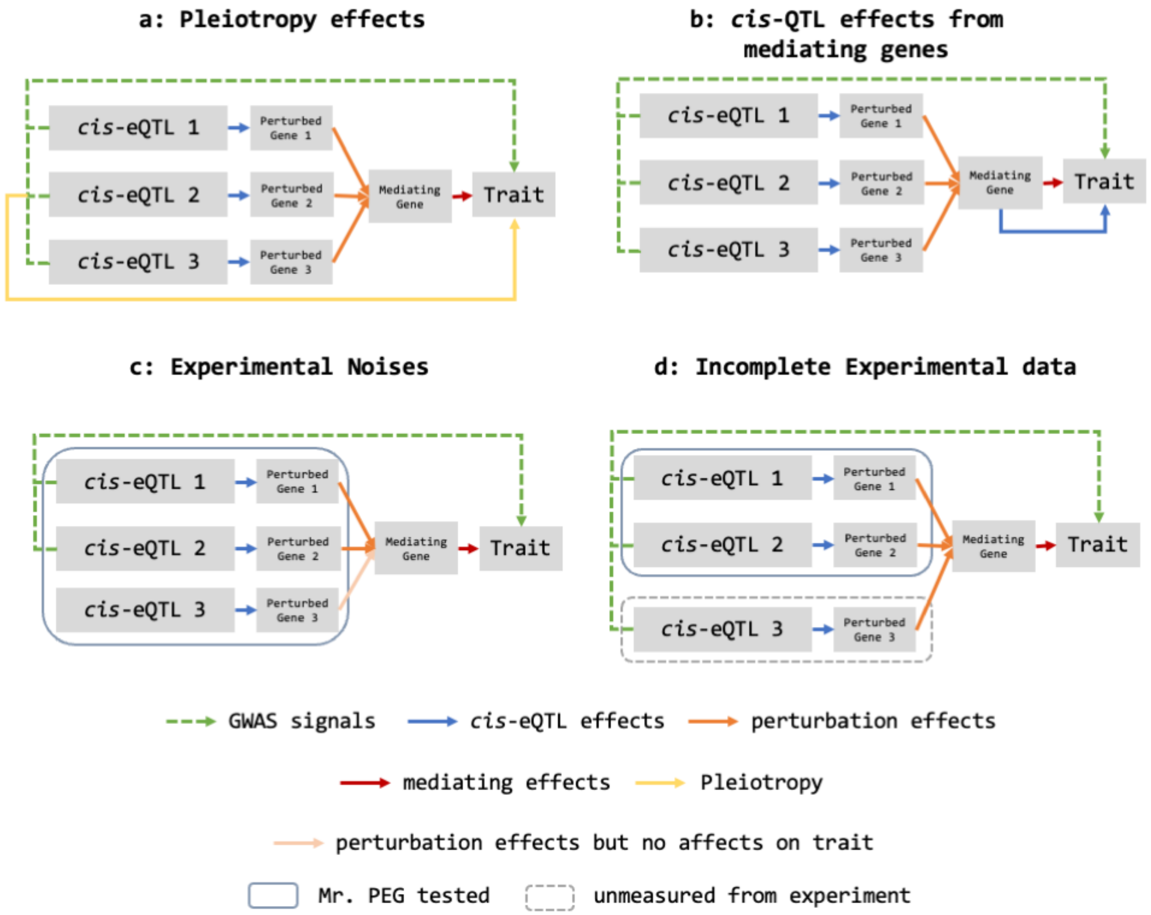


**Supplementary Figure 3: Model misspecification scenarios.**

(a) *cis*-eQTLs of perturbed genes (pGEs) exhibit pleiotropic effects on complex traits. (b) Expression of mediating genes (mGEs) is influenced both by perturbed genes and by their own *cis*-eQTLs, thereby affecting downstream complex traits. (c) Additional perturbed genes are included in the experiment and are thus included in the Mr. PEG inference framework. (d) Some perturbed genes that contribute to complex traits are not included in the experiment and are therefore missed by the Mr. PEG inference framework.

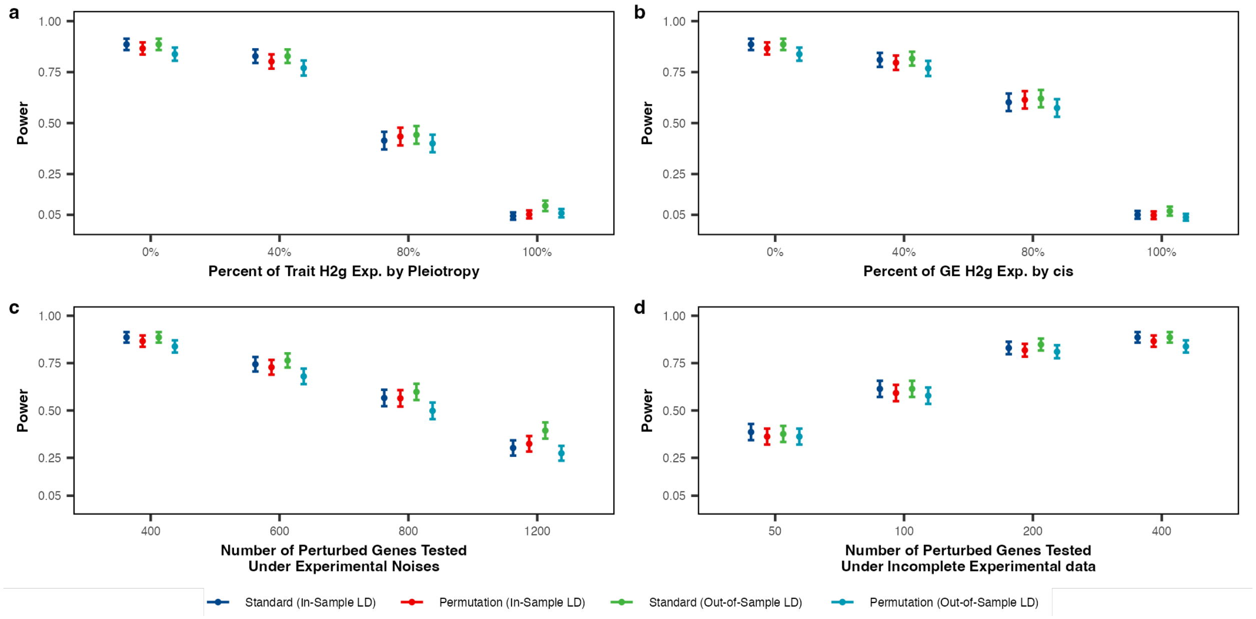


**Supplementary Figure 4: Mr. PEG remains robust in power under model misspecification scenarios.**

(a) Percentage of trait heritability explained by cis-eQTL pleiotropic effects, where 100% means no perturbation effects contribute to the complex trait. (b) Percentage of gene expression heritability of the mediating gene explained by its own cis-eQTLs, where 100% means no perturbation effects contribute to the mediating gene. (c) Number of perturbed genes tested in the inference when 400 genes contribute to the complex trait; label 400 indicates the baseline where no additional perturbed genes are included. (d) Number of contributing perturbed genes missed in the inference; label 400 indicates the baseline where all contributing perturbed genes are included. We compared four versions of Mr. PEG: (1) using standard MLE with its analytically derived standard error ("Standard"), (2) using MLE with standard error estimated from permutation tests ("Permutation"), and (3–4) using either an in-sample or an out-of-sample LD matrix (**Methods**). Each parameter setting was evaluated using 500 independent replicates. Power was calculated as the proportion of replicates (out of 500) with a P value < 0.05 under the alternative scenario. Unless otherwise specified, the default parameters were: GWAS sample size = 100,000; *cis*-eQTL sample size = 200; number of *cis*-eQTLs = 1; number of perturbed genes contributing to the complex trait = 400; number of perturbed genes tested in inference = 400; per-gene trait heritability = $3 \times 10^{-4}$; no pleiotropic effects; and the mediating gene has no *cis*-eQTLs. The points are the mean across simulations, and the error bars are their corresponding 95% confidence intervals.

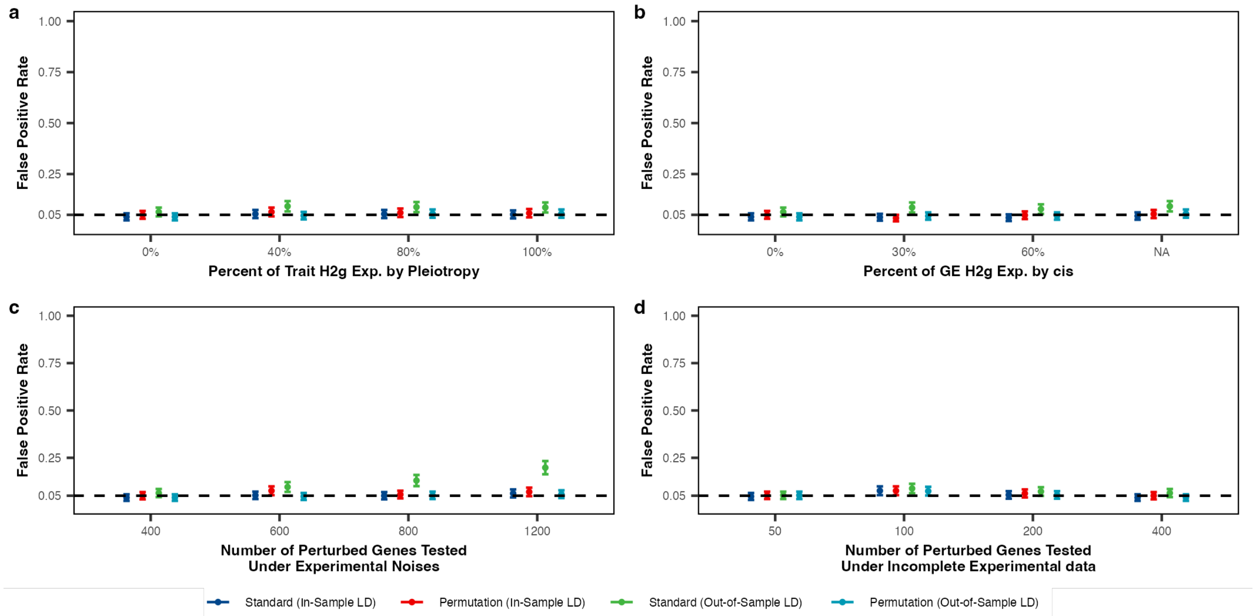


**Supplementary Figure 5: Mr. PEG controls the false positive rate under model misspecification scenarios.**

(A) Percentage of trait heritability explained by cis-eQTL pleiotropic effects, where 100% means no perturbation effects contribute to the complex trait. (B) Percentage of gene expression heritability of the mediating gene explained by its own cis-eQTLs, where 100% means no perturbation effects contribute to the mediating gene. (C) Number of perturbed genes tested in the inference when 400 genes contribute to the complex trait; label 400 indicates the baseline where no additional perturbed genes are included. (D) Number of contributing perturbed genes missed in the inference; label 400 indicates the baseline where all contributing perturbed genes are included. We compared four versions of Mr. PEG: (1) using standard MLE with its analytically derived standard error ("Standard"), (2) using MLE with standard error estimated from permutation tests ("Permutation"), and (3–4) using either an in-sample or an out-of-sample LD matrix (see **Methods**). Each parameter setting was evaluated using 500 independent replicates. The false positive rate was calculated as the proportion of replicates (out of 500) with a P value < 0.05 under the null scenario. Unless otherwise specified, the default parameters were: GWAS sample size = 100,000; *cis*-eQTL sample size = 200; number of *cis*-eQTLs = 1; number of perturbed genes contributing to the complex trait = 400; number of perturbed genes tested in inference = 400; trait heritability = $3 \times 10^{-4}$; no pleiotropic effects; and the mediating gene has no *cis*-eQTLs. The points are the mean across simulations, and the error bars are their corresponding 95% confidence intervals.

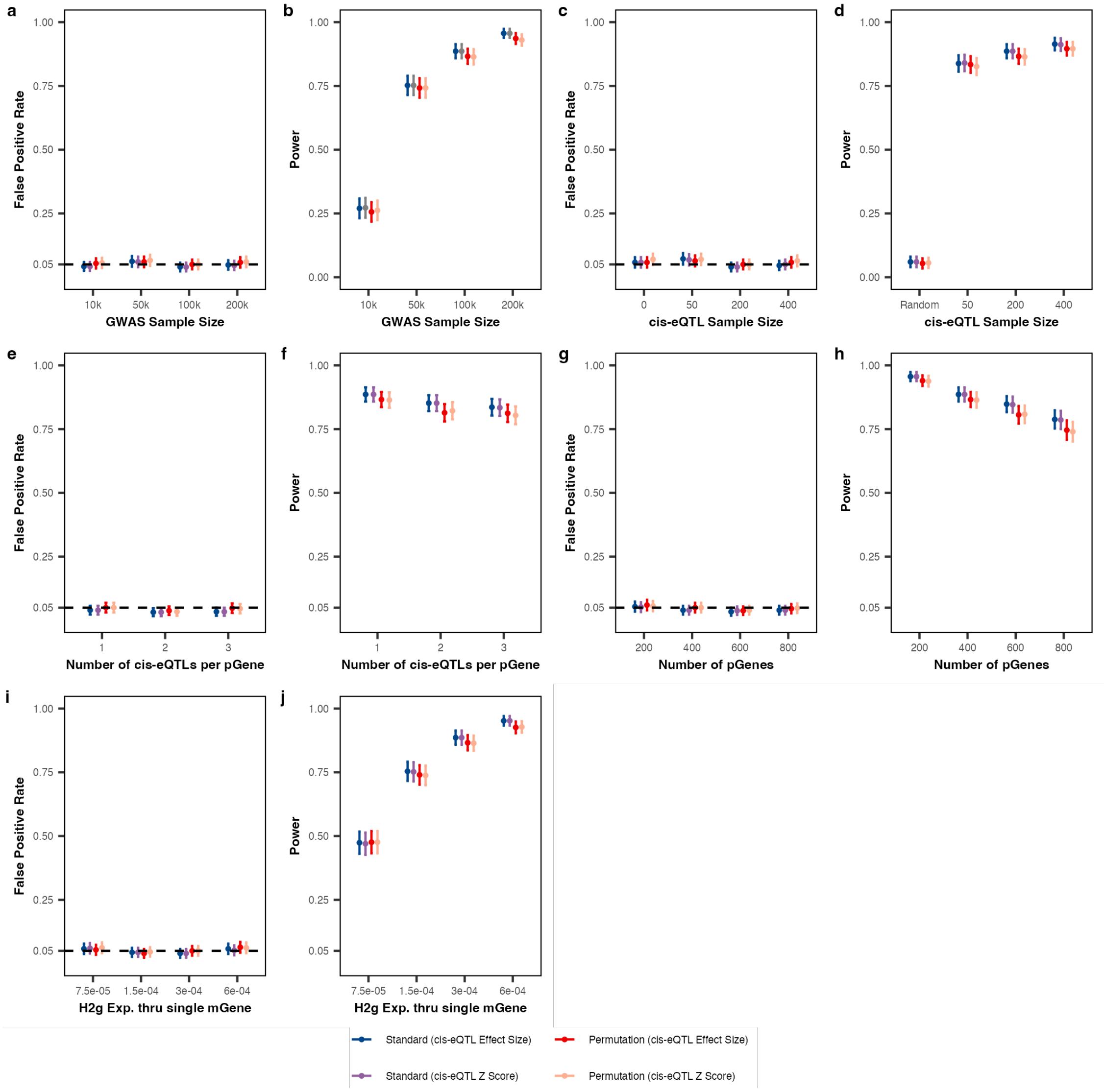


**Supplementary Figure 6: No difference in Mr.PEG framework results between using *cis*-eQTL effect sizes and Z-scores.**

(a, c, e, g, i) False positive rate; we simulated scenarios under the null (no perturbation effects; "Null" in **Fig. 1**) (b, d, f, h, j) Power, which was calculated after Bonferroni correction; we simulated scenarios under the alternative (with perturbation effects; "Alternative" in **Fig. 1**). We compared four versions of Mr. PEG: (1) using standard MLE with its analytically derived standard error ("Standard"), (2) using MLE with standard error estimated from permutation tests ("Permutation"), and (3–4) using either cis-eQTL Effect Size or cis-eQTL Z score (**Methods**). In-sample LD matrix was used in the matrix. Each parameter setting was evaluated using 500 independent replicates. The false positive rate was calculated as the proportion of replicates (out of 500) with a P value < 0.05 under the null scenario. Power was calculated as the proportion of replicates (out of 500) with a P value < 0.05 under the alternative scenario. Unless otherwise specified, the default parameters were: GWAS sample size = 100,000; *cis*-eQTL sample size = 200; number of *cis*-eQTLs = 1; number of perturbed genes contributing to the complex trait = 400; number of perturbed genes tested in inference = 400; per-gene trait heritability = $3 \times 10^{-4}$; no pleiotropic effects; and the mediating gene has no *cis*-eQTLs. The points are the mean across simulations, and the error bars are their corresponding 95% confidence intervals.

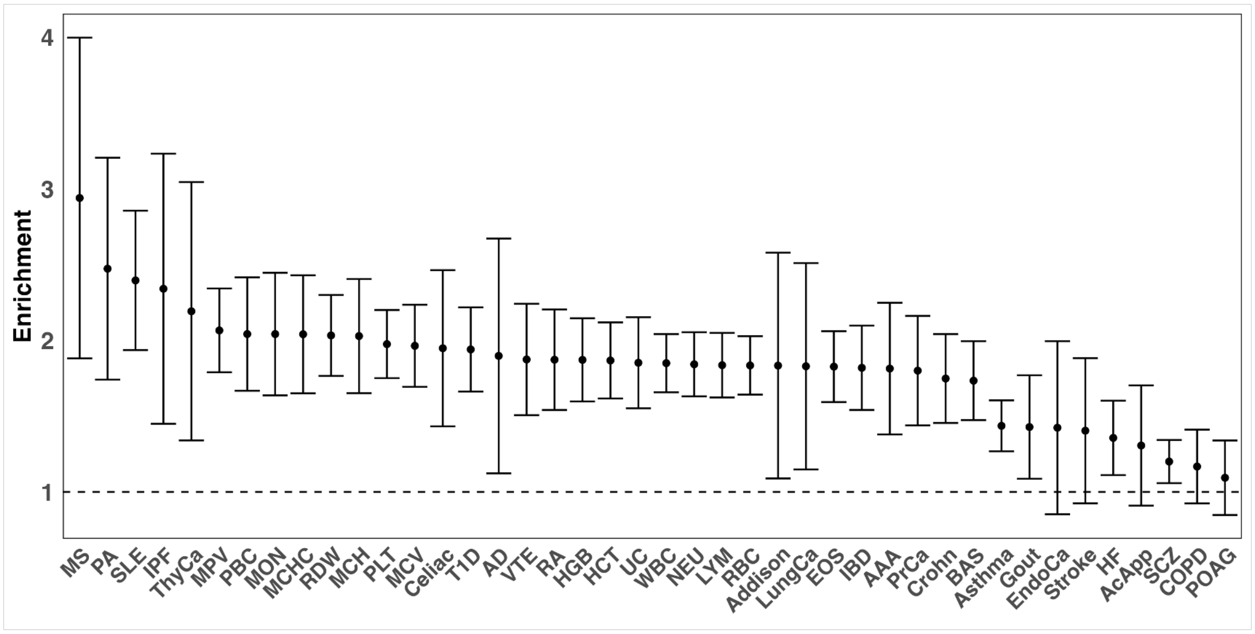


**Supplementary Figure 7: Top 500 most highly expressed genes from Yao et al. show significant heritability enrichment across complex traits.**

We selected the top 500 most highly expressed genes in control cells from the Perturb-seq experiment by Yao et al. We used this gene set to create a binary genomic annotation and ran stratified LD score regression (LDSC), finding that these genes are significantly enriched for the heritability of the complex traits analyzed. The points are the enrichment for each trait and the error bars are their corresponding 95% confidence interval.

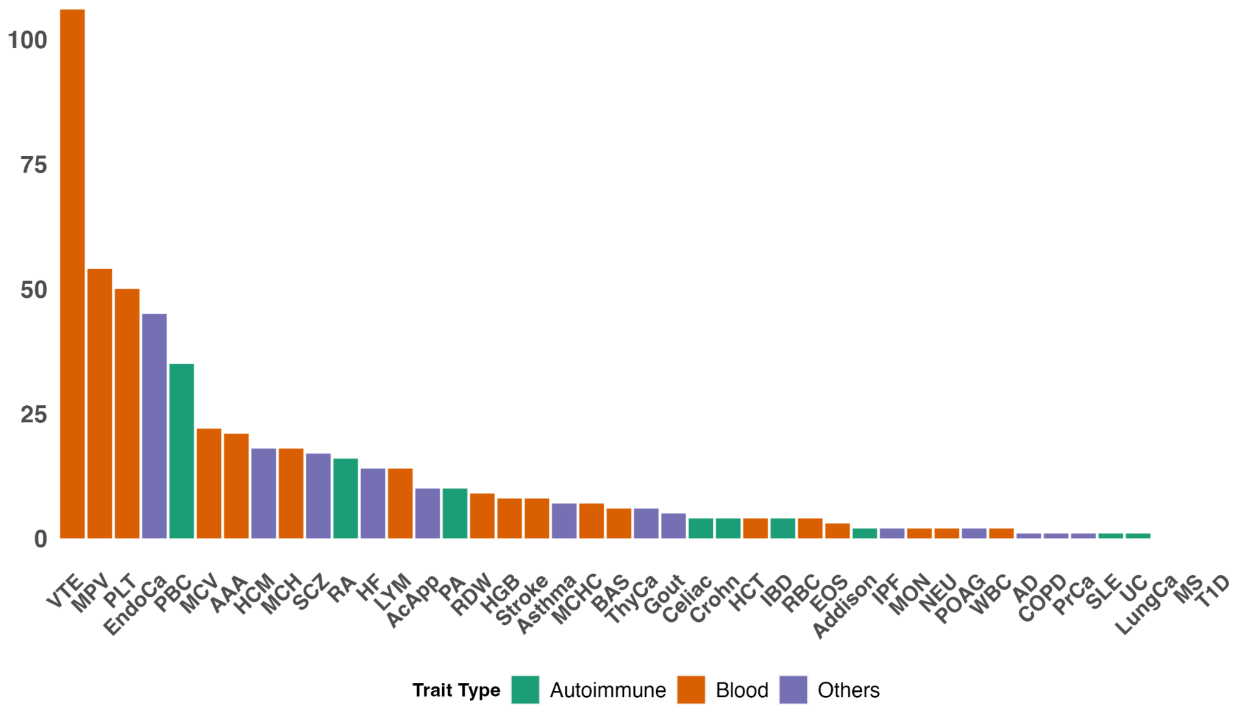


**Supplementary Figure 8: Mr. PEG identifies 543 significant genes across 40 complex traits.**

This result aggregates analyses using two types of Perturb-seq effect size matrices: the full matrix and a sparse matrix containing the top 1% of absolute effect sizes. Statistical significance level is 0.05 based on adjusted P value using Bonferroni correction with n = 23,000.

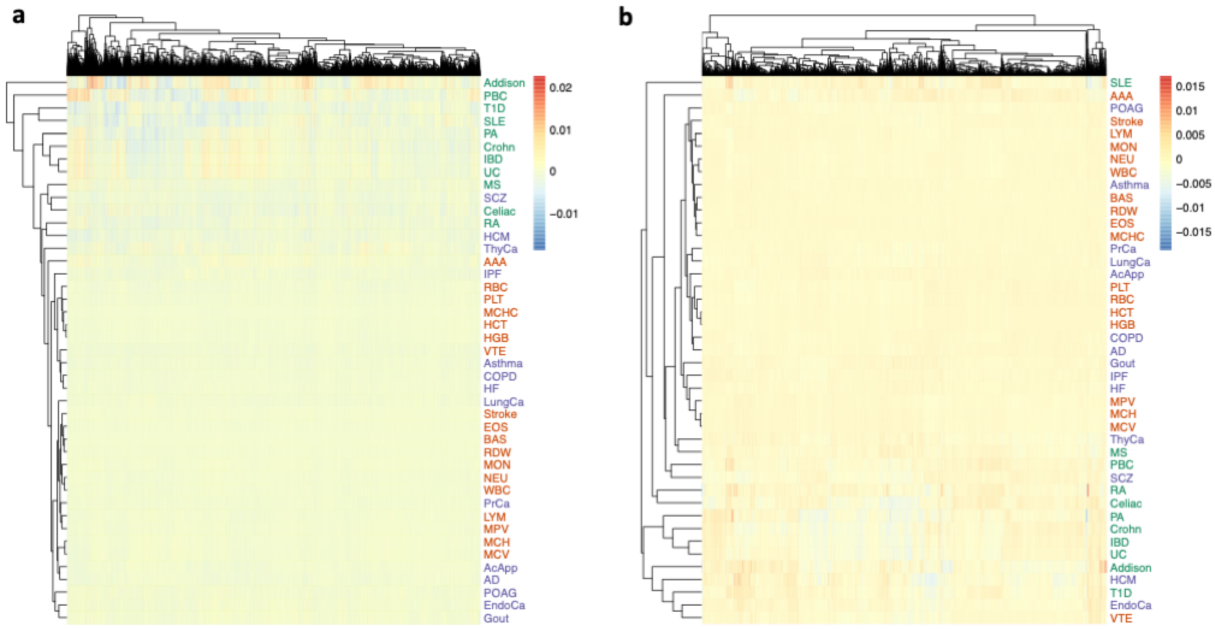


**Supplementary Figure 9: Mr. PEG effect sizes are larger for autoimmune diseases compared to other complex traits.**

Heatmap of Mr. PEG effect sizes (rather than z statistics), using (a) the full Perturb-seq effect size matrix and (b) a sparse matrix retaining only the top 1% of absolute effect sizes. Green indicates autoimmune diseases, orange indicates blood-related traits, and purple indicates other complex traits. For the full matrix, the absolute Mr. PEG effect sizes for autoimmune diseases are 3.9-fold and 2.4-fold greater than those for blood-related traits and other complex traits, respectively. For the sparse matrix, the absolute Mr. PEG effect sizes for autoimmune diseases are 4.7-fold and 2.8-fold greater than those for blood-related traits and other complex traits, respectively.

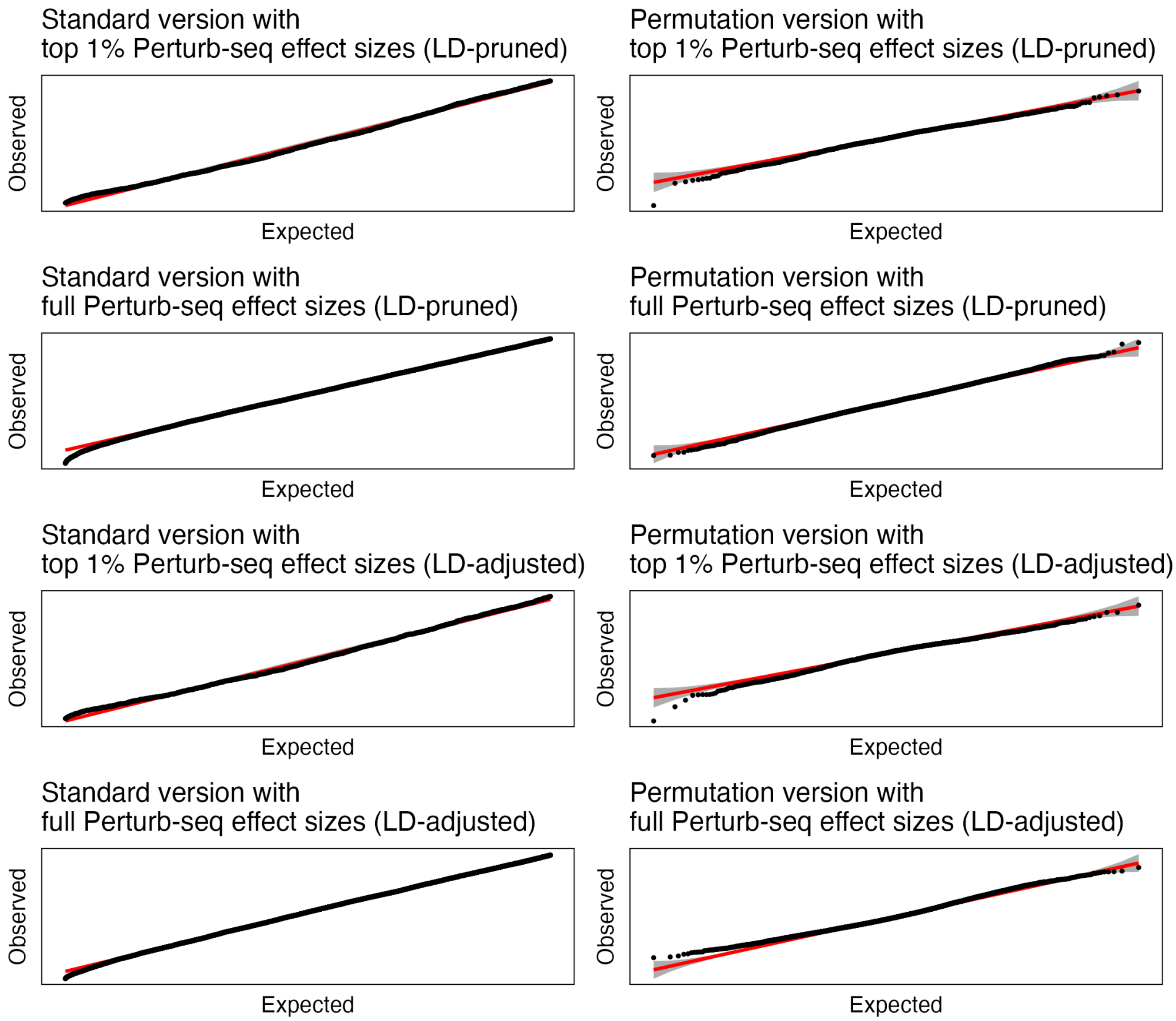


**Supplementary Figure 10: Mr. PEG produces well-calibrated results using shuffled GWAS effect sizes and real data for *cis*-eQTL and Perturb-seq.**

We evaluated multiple versions of Mr. PEG. Specifically, we ran the Standard version, which uses maximum likelihood estimation (MLE) with analytically derived standard errors, and the Permutation version, which uses MLE with permutation-based standard errors. We tested two types of Perturb-seq effect size matrices: the full matrix and a sparse matrix retaining only the top 1% of absolute effect sizes. Additionally, we assessed models with different linkage disequilibrium (LD) adjustments: one using out-of-sample LD from the 1000 Genomes Project and another applying LD pruning. For visualization, the Standard version's Q-Q plot compares p-values to a uniform distribution, while the Permutation version compares Z-scores to a standard normal distribution.

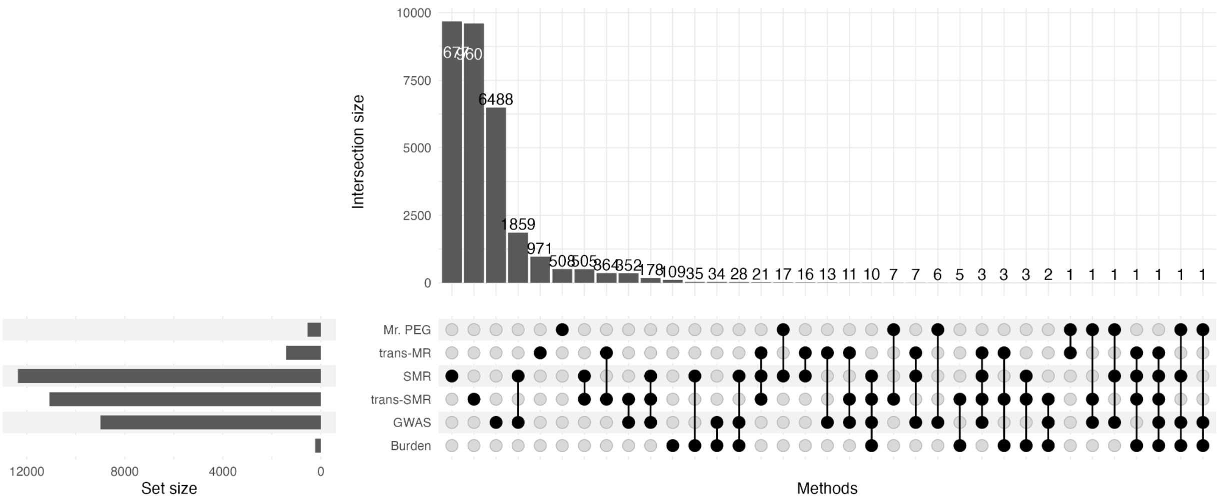


**Supplementary Figure 11: The majority of Mr. PEG genes (93%) are uniquely identified compared to other methods.**

Upset plot showing the overlap of significant genes identified by each method. Each count represents a unique trait–gene pair.

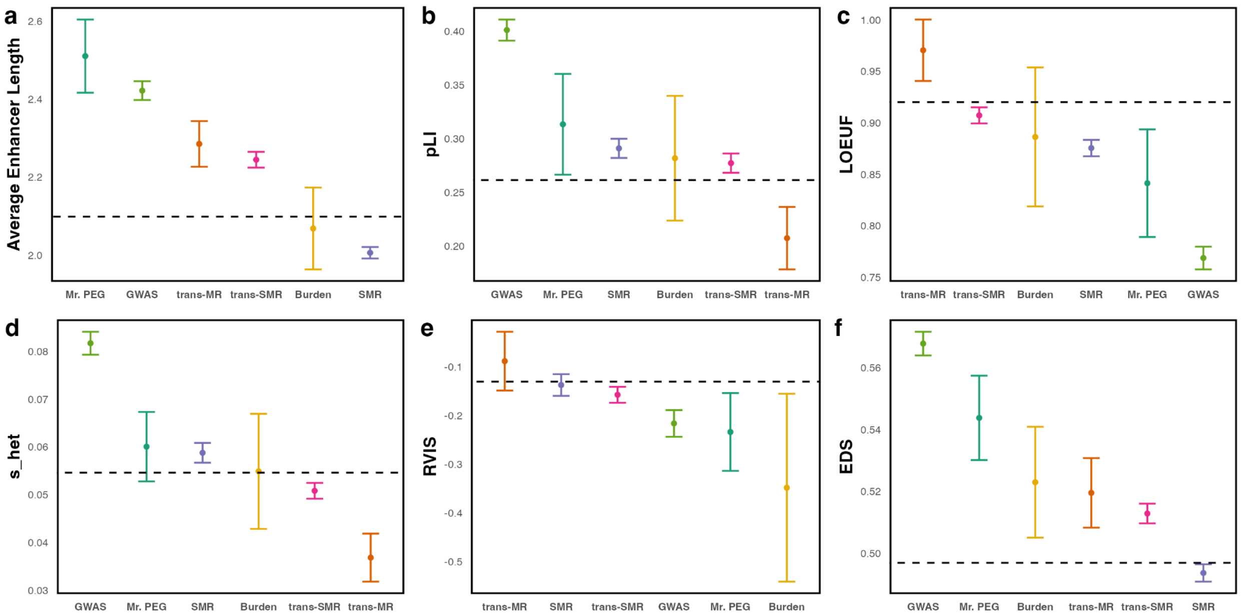


**Supplementary Figure 12: Mr. PEG genes exhibit longer lengths across enhancers and are more constrained compared to other methods.**

Annotations curated by Mostafavi et al. (a) Average lengths across enhancers from Nasser et al. (b-f) different metrics that correlate with loss-of-function intolerance. The standard error is calculated using bootstrap. The points are the mean across simulations, and the error bars are their corresponding 95% confidence intervals.

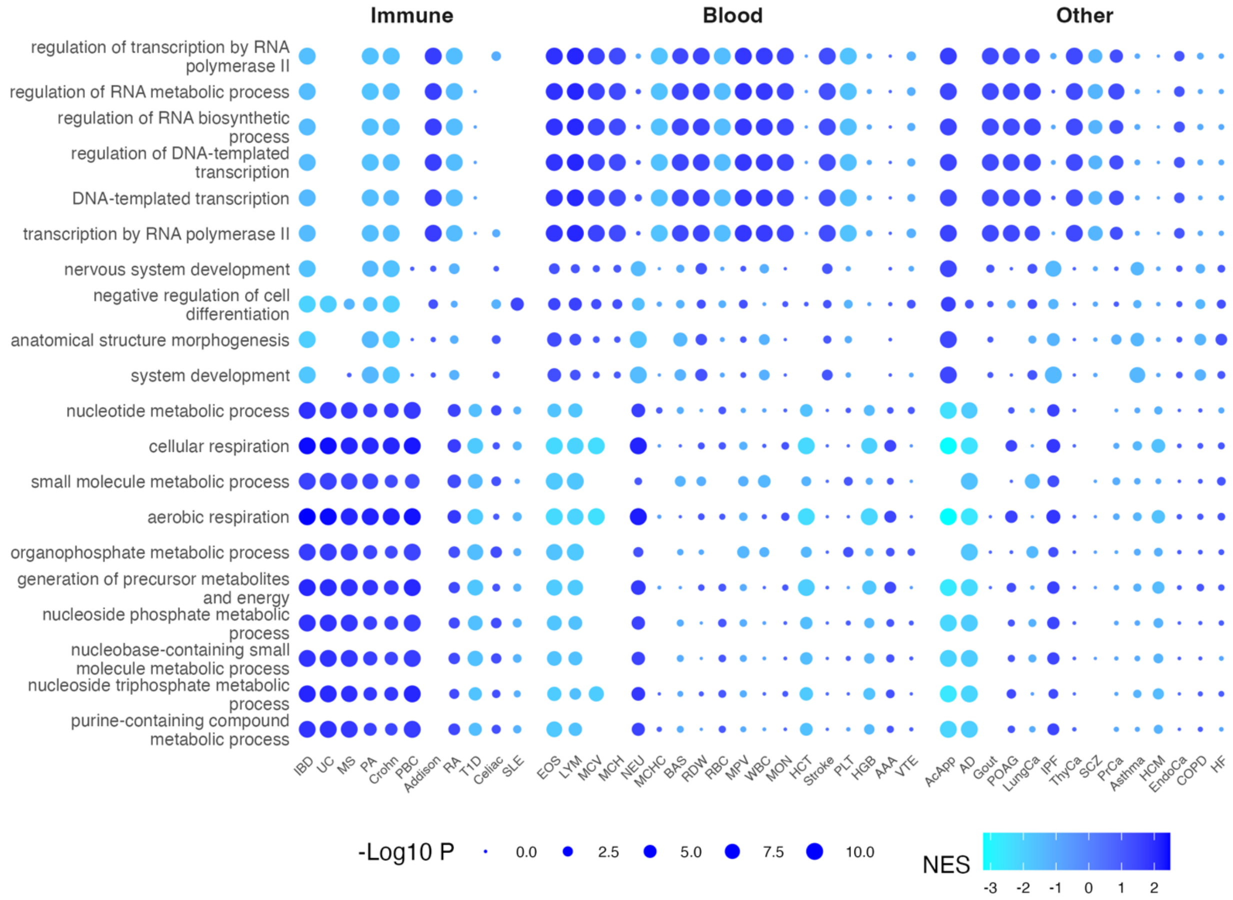


**Supplementary Figure 13: Mr. PEG genes in immune-related diseases where higher expression was linked to increased disease risk were mainly enriched in transcriptional regulatory processes**

We ranked genes by their nominal Mr. PEG statistics and performed GSEA. A negative normalized enrichment score indicates gene sets in which higher expression is associated with increased trait levels or risk, whereas a positive score indicates the opposite. Shown are the ten most negative and ten most positive normalized enrichment scores across immune related traits, presented from increased expression at the top to decreased expression at the bottom. The corresponding results for other complex traits and blood-related traits are shown on the right, clearly illustrating regulatory heterogeneity between immune-related diseases and those related to blood and other traits.

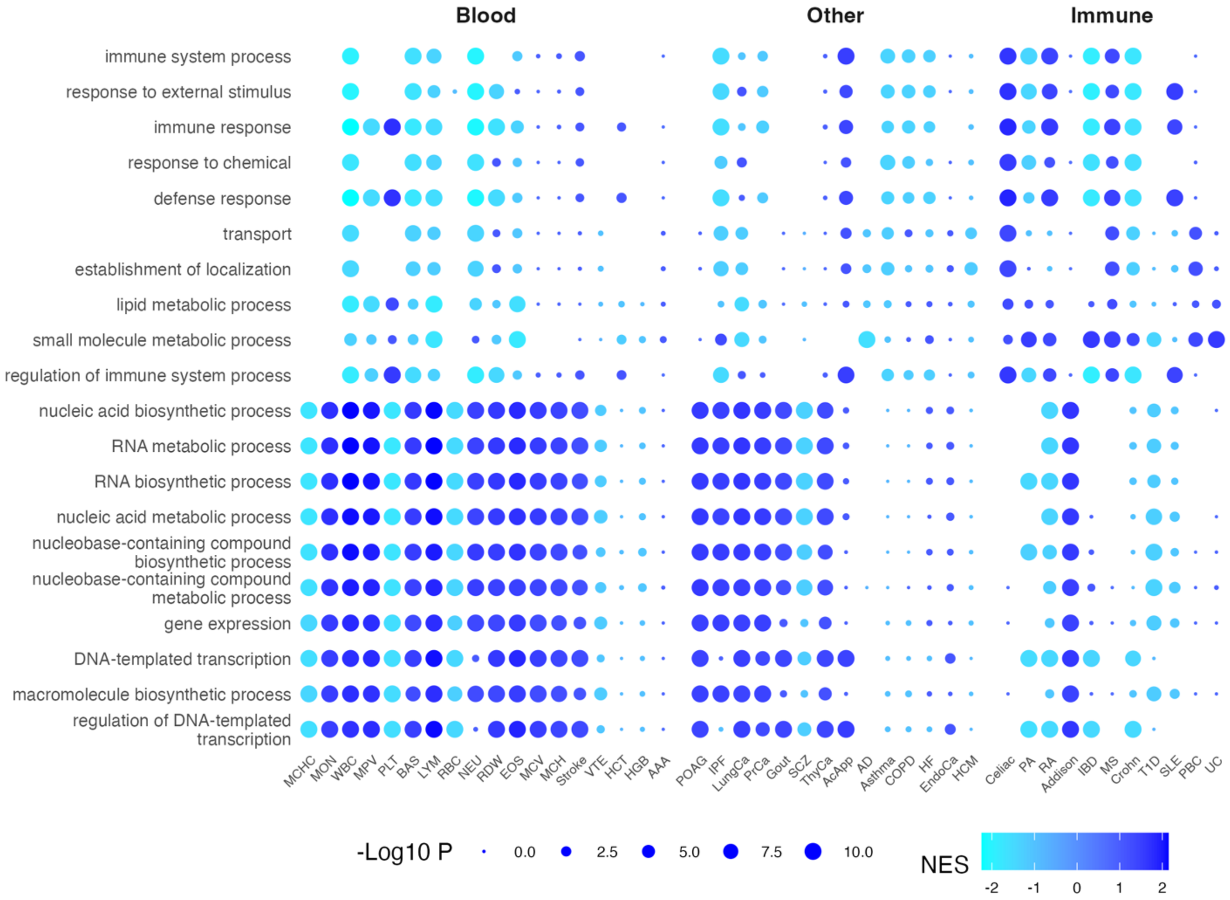


**Supplementary Figure 14: Mr. PEG genes in blood-related traits where lower expression was linked to increased disease risk were mainly enriched in transcriptional regulatory processes and nucleic acid metabolic processes.**

We ranked genes by their nominal Mr. PEG statistics and performed GSEA. A negative normalized enrichment score indicates gene sets in which higher expression is associated with increased trait levels or risk, whereas a positive score indicates the opposite. Shown are the ten most negative and ten most positive normalized enrichment scores across immune blood-related traits, presented from increased expression at the top to decreased expression at the bottom. The corresponding results for immune-related complex traits and other complex traits are shown on the right, clearly illustrating regulatory heterogeneity between immune-related diseases and those related to blood and other traits.

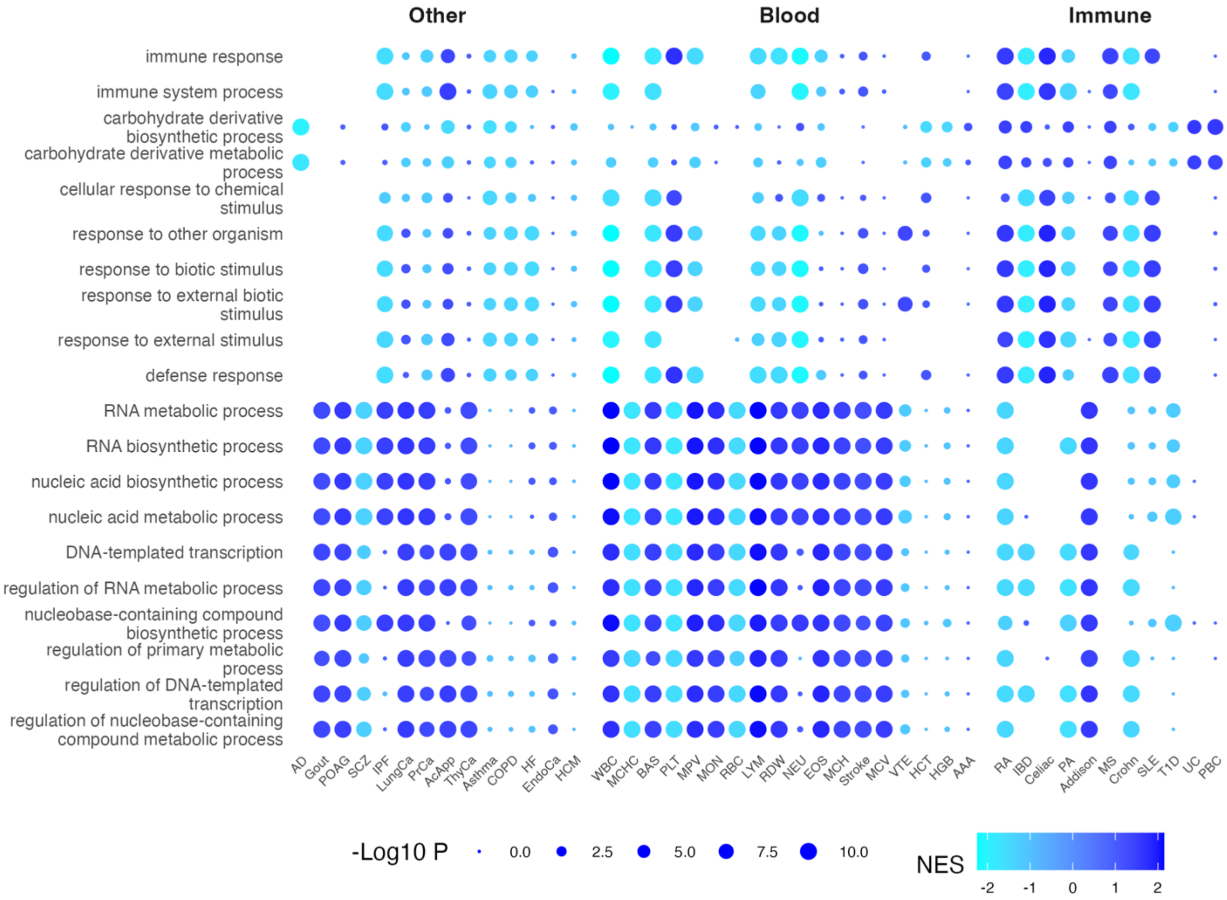


**Supplementary Figure 15: Mr. PEG genes in other complex traits where higher expression was linked to increased disease risk were mainly enriched in immune and defense activation pathways.**

We ranked genes by their nominal Mr. PEG statistics and performed GSEA. A negative normalized enrichment score indicates gene sets in which higher expression is associated with increased trait levels or risk, whereas a positive score indicates the opposite. Shown are the ten most negative and ten most positive normalized enrichment scores across other complex traits, presented from increased expression at the top to decreased expression at the bottom. The corresponding results for blood-related traits and immune-related diseases are shown on the right, clearly illustrating regulatory heterogeneity between immune-related diseases and those related to blood and other traits.

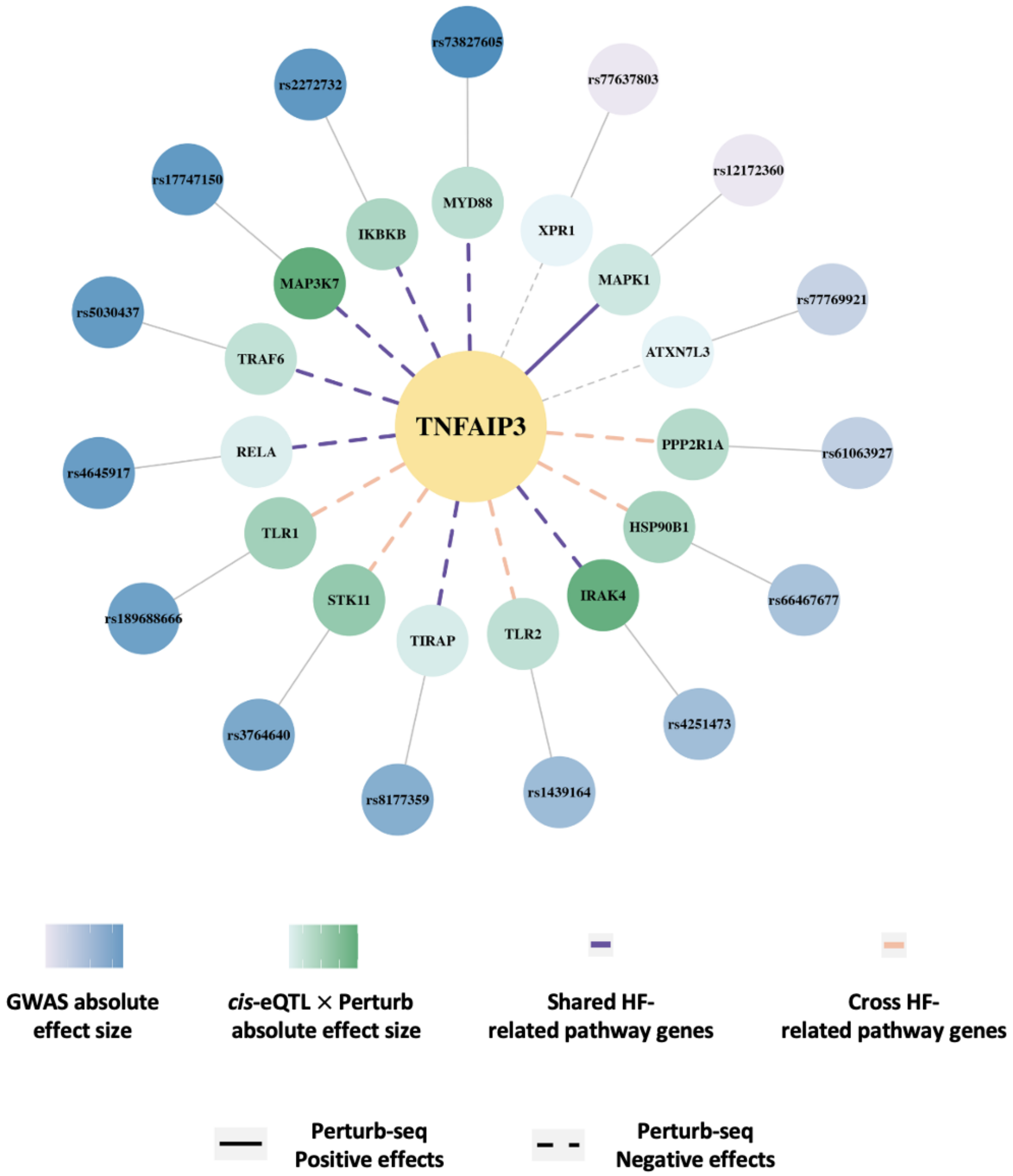


#### Supplementary Figure 16: Mr. PEG identifies *TNFAIP3* for heart failure.

Perturbed genes of *TNFAIP3* are shown in green, with shading indicating the product of *cis*-eQTL effect size and Perturb-seq effect size. Perturbed genes are defined as those with effect sizes in the top 1% of the original Perturb-seq matrix. The top *cis*-eQTL for each perturbed gene (i.e., the most significant) is shown in blue, with shading representing the absolute GWAS effect size. Genes are ordered by the GWAS effect size of their top *cis*-eQTL. Colored lines (purple or orange) connect genes appearing in KEGG pathways related to heart failure: purple indicates both genes are in the same pathway, while orange indicates they are in different but closely associated pathways. Solid lines represent positive Perturb-seq effects; dashed lines indicate negative effects. None of the genes are SMR significant, and none of the cis-eQTLs are GWAS significant.

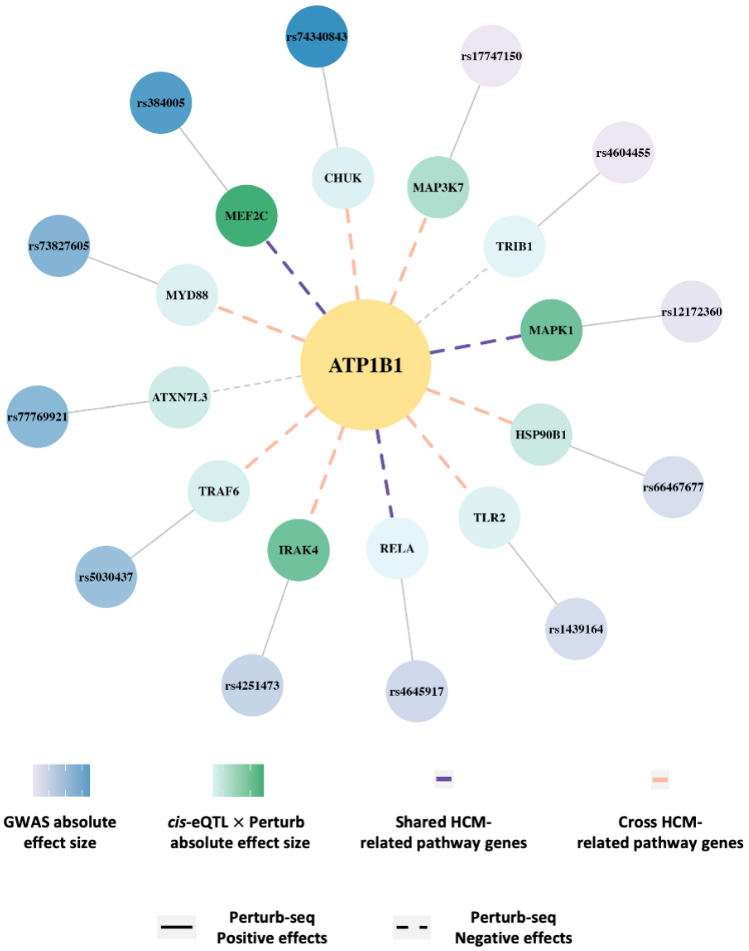


**Supplementary Figure 17: Mr. PEG identifies *ATP1B1* for hypertrophic cardiomyopathy.**

Perturbed genes of *ATP1B1* are shown in green, with shading indicating the product of *cis*-eQTL effect size and Perturb-seq effect size. Perturbed genes are defined as those with effect sizes in the top 1% of the original Perturb-seq matrix. The top *cis*-eQTL for each perturbed gene (i.e., the most significant) is shown in blue, with shading representing the absolute GWAS effect size. Genes are ordered by the GWAS effect size of their top *cis*-eQTL. Colored lines (purple or orange) connect genes appearing in KEGG pathways related to hypertrophic cardiomyopathy (HCM: purple indicates both genes are in the same pathway, while orange indicates they are in different but closely associated pathways. Solid lines represent positive Perturb-seq effects; dashed lines indicate negative effects. None of the genes are SMR significant, and none of the cis-eQTLs are GWAS significant.
